# Patterns and Clinical Outcomes of Physical Activity and Sedentary Behavior Across 20 Million Days of Wearable Monitoring in U.S. Adults

**DOI:** 10.64898/2026.02.11.26346079

**Authors:** Arash A. Nargesi, Valentina D’Souza, Tal Shnitzer, Andri Kadaifciu, Aurora Cremer, Sean Joseph Jurgens, Nakia Mack, Ryan Lupi, Romuladus Azuine, Geoffrey S. Ginsburg, Chris Lunt, Christopher D. Anderson, Sam Friedman, Patrick T. Ellinor, Mahnaz Maddah

**Affiliations:** Data Sciences Platform, Broad Institute of MIT and Harvard, Cambridge, Massachusetts, USA; Division of Cardiovascular Medicine, Brigham and Women’s Hospital, Boston, Massachusetts, USA; Clinical Informatics, Mass General Brigham, Somerville, Massachusetts, USA; Cardiovascular Disease Initiative, Broad Institute of MIT and Harvard, Cambridge, Massachusetts, USA; All of Us Research Program, National Institutes of Health, Bethesda, MD, USA; McCance Center for Brain Health, Massachusetts General Hospital, Boston, USA; Department of Neurology, Brigham and Women’s Hospital, Boston, USA; Cardiovascular Research Center, Massachusetts General Hospital, Boston, Massachusetts, USA

**Keywords:** Physical activity, step count, sedentary behavior, wearables, digital health

## Abstract

**Background:** Physical activity is a key modifiable determinant of health, yet current guidelines primarily emphasize moderate-to-vigorous physical activity and provide limited guidance on other actionable dimensions such as daily step count and sedentary time. The growing availability of wearables enables high-resolution measurement of these metrics and assessment of their associations with clinical outcomes.

**Methods:** Minute-level wearable data from adult participants of *All of Us* between 2015-2023 were used to calculate step count and sedentary time. Patterns of step count and sedentary behavior were assessed across temporal scales, U.S. states, and sociodemographic groups. Phenome-wide association (PheWAS) and dose-response analyses were conducted to evaluate associations with incident disease.

**Results:** Among 50,300 participants (68% female, 71% White, median age 55 years), 19,845,612 person-days of wearable monitoring over a median follow-up of 13 months were included in this analysis. Step counts peaked at midday with weekly highs on Saturdays and seasonal highs in May, while sedentary time was highest in midafternoon and winter. Higher step counts and lower sedentary time were observed among males, individuals with higher income and education, and residents of the Northeast, upper Midwest, and West Coast. In PheWAS (n=31,446), higher step count and lower sedentary time were associated with lower risk of multiple diseases, including obesity, cardiometabolic risk factors, and mood and anxiety disorders. Dose response analyses demonstrated heterogeneous relationships across disease groups, with cardiovascular benefits plateauing beyond 9,000-10,000 steps/day and below 600-640 minutes/day of sedentary time.

**Conclusions:** In this nationwide cohort of U.S. adults, wearable-derived physical activity and sedentary behavior showed distinct temporal and geographic patterns with significant disease-specific associations with clinical outcomes. These findings support the use of longitudinal high-resolution wearable data for advancing personalized guidelines of physical activity and sedentary behavior.

## Introduction

Physical activity is widely recognized as one of the most effective and modifiable determinants of health^1^. Clinical guidelines from the World Health Organization (WHO) and American Heart Association (AHA) recommend adults to engage in at least 150 minutes of moderate or 75 minutes of vigorous aerobic activity per week^2,3^. The Physical Activity Guidelines for Americans also recommend reducing sedentary behavior as a key strategy for preventing chronic diseases^4^. Despite the robust evidence supporting the role of physical activity in human health, current guidelines mostly focus on moderate and vigorous physical activity (MVPA), while target levels for other dimensions of physical activity, such as daily step count and sedentary time have not been clearly established.

Physical activity encompasses diverse combinations of movement intensity and duration, and its patterns among individuals are highly variable and yet poorly characterized. A key barrier to developing universally applicable guidelines has been the limited scope and short follow-up of previous studies in this domain. Large-scale studies, including community cohorts such as the UK biobank^1,5^ and nationally representative surveys such as National Health and Nutrition Examination Survey (NHANES)^6^, were limited to 1-2 weeks of monitoring. Nevertheless, the growing availability of wearable technology has significantly enhanced the feasibility and accuracy of tracking physical activity at scale. Among the contemporary cohorts, the *All of Us* (AoU) program from the US National Institutes of Health (NIH) provides a unique opportunity to access longitudinal wearable data collected from a broad sample of individuals across the US. The long-term follow-up and linkage of wearable data with electronic health records (EHRs) facilitate the assessment of longitudinal patterns of physical activity^7^. Furthermore, understanding the exact relationship of step count and sedentary time with future risk of disease is essential to inform personalized and actionable guidance on physical activity and complement existing recommendations based on MVPA alone.

In this study, we used minute-level wearable data from adult participants in the AoU Program to characterize temporal, geographic, and sociodemographic patterns of daily step count and sedentary time across the US. We further evaluated their associations with incident disease using phenome-wide and dose–response analyses. By linking high-resolution wearable data with longitudinal clinical outcomes, this study provides evidence relevant to the use of wearable-derived physical activity metrics in population health and preventive medicine. In addition to describing population-level activity patterns, we aimed to identify disease-specific step count and sedentary time thresholds associated with a lower risk of incident disease.

## Methods

### Study Design and Participants

This was a prospective cohort study using longitudinal wearable data and EHRs from the National Institutes of Health AoU Program^8,9^. Details of study protocols have been previously published^10,11^. We used registered and controlled tier datasets (Version 8, ID numbers R2024Q3R3 and C2024Q3R4) which include data from participants enrolled between 2015 and 2023. Adults aged 18 years or older with available wearable data were eligible for inclusion. The analytic cohort was derived as shown in the CONSORT flow diagram (**Figure 1**). Demographic and survey data were collected at enrollment, and EHRs were linked from participating health systems. All participants provided informed consent at enrollment, including consent to share wearable data. This study was conducted in accordance with the Strengthening the Reporting of Observational Studies in Epidemiology (STROBE) reporting guideline.

**Figure 1.**
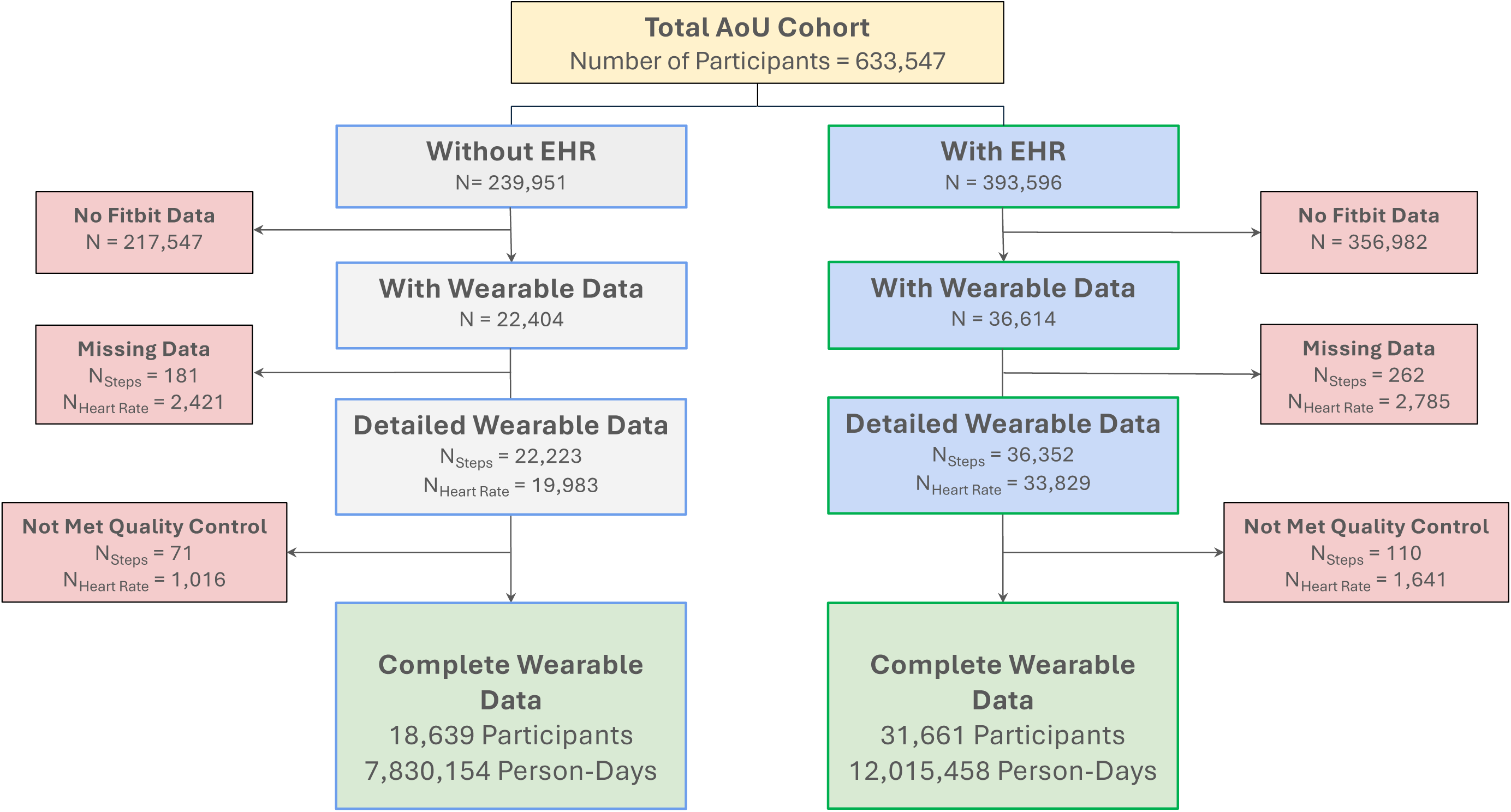
Study Population. The CONSORT diagram illustrates the inclusion and exclusion of participants from the All of Us Research Program. Wearable data, including step counts and heart rates, that passed quality control were included in the final analysis. Abbreviations: EHR, Electronic Health Records.

### Wearable Data

Wearable data in the AoU Program were collected using Fitbit devices. Participants were enrolled through 2 pathways: (1) Bring Your Own Device (BYOD), in which participants linked a personal Fitbit account to the All of Us participant portal, and (2) the Wearable Enhancing All of Us Research (WEAR) study, in which Fitbit devices (Charge 4 or Versa 3) were provided to participants at no cost. Details of enrollment procedures, device distribution, and data collection have been reported previously^12,13^.

Fitbit devices continuously measure movement and physiologic signals, including accelerometer-based activity and photoplethysmography-derived heart rate, which are processed using proprietary algorithms to generate activity metrics. AoU provides second- or minute-level step counts and daily summaries of physical activity across intensity zones to the researchers, including total daily minutes spent at sedentary, light, moderate, vigorous, and peak activity zones.

### Data Preprocessing

We used minute-level step count and heart rate to assess the patterns of physical activity and sedentary behavior in the study population. The raw minute-level data are noisy and require substantial preprocessing to ensure adequate quality for downstream analysis. We converted step count and heart rate data into numerical minute-level vectors, identified and excluded non-wear times, removed outliers based on physiological ranges of heart rate and step count, and excluded records with high missingness. Fitbit trackers do not provide data on sedentary behavior at minute-level scale. To comprehensively assess the patterns of physical activity, we estimated minute-level sedentary times from the accelerometer and heart rate data. Procedural methods, quality control, and calculations of sedentary time are described in **Supplement** (**Method description**).

### Study Outcomes and Covariates

We assessed temporal and geographic patterns of step count and sedentary time, evaluated their associations with the incidence of clinical phenotypes in a PheWAS analysis, and conducted dose-response analyses to identify potential thresholds of step count and sedentary time associated with lower risk of incident disease.

Sociodemographic data, including age, sex, self-reported race, education, income, employment, and the state of residence were collected at enrollment. Clinical characteristics were derived from the EHRs using standardized coding systems. Corresponding codes for clinical phenotypes were grouped using phecodeX, a phenotyping tool that adds granularity and improves the hierarchical structure of under-represented phenotypes^14^. Using phecodeX definitions, we performed a PheWAS analysis to examine the association of step count and sedentary time with incident cardiovascular, metabolic, musculoskeletal, gastrointestinal, respiratory, neoplastic, neurological, and mental diseases.

We identified incident events as the first appearance of disease diagnosis codes in the EHRs after applying a blanking period of 30 days from the last wearable monitoring. All wearable data over a maximum time horizon of 3 years preceding the incident event were included in the analysis. The 3-year limit was applied to minimize the influence of long-term behavioral changes that could weaken the temporal relevance between wearable-derived predictors and disease outcomes. In all prospective analyses, data from the control group were restricted to wearable monitoring between 2015-2022 to temporally align with the incident group. All data beyond the incidence date or the last clinical encounter were censored.

### Statistical Analysis

*a. Descriptive Analyses*: Temporal patterns of daily step count and sedentary time were examined across multiple time scales, including within-day, weekly, monthly, and yearly intervals, as well as over an extended follow-up period of up to 5 years. For each participant, average daily profiles were first computed for each metric and subsequently aggregated to generate population-level estimates, with equal weighting assigned to all participants. In sensitivity analyses, we examined daily and weekly trends of step count across sociodemographic subgroups and within each enrollment pathway (BYOD vs. WEAR). As sedentary time is calculated in minutes, its daily distribution represents the proportion of participants engaged in such activities at each minute point in time. To ensure consistency and minimize the confounding effect of early dropouts, yearly analyses were restricted to individuals with available data for at least 12 contiguous months and multi-year analyses to participants with complete five-year follow-ups. For participants with non-contiguous periods of monitoring and gaps exceeding 3 months, only the longest continuous data were used in yearly and multi-year analyses. To assess geographical variations of physical activity, each metric was aggregated at the state level and visualized in choropleth maps. Similar to our analysis of temporal patterns, equal weights were given to all participants within each state.
*b. PheWAS*: We also examined the association of step count and sedentary time with incident disease conditions using phenotypic codes described above and identified the top five associations in each disease category based on the numerical p-values. These analyses were restricted to individuals with available EHRs (n=31,446). Each physical activity metric was tested in multivariable Cox regression models adjusted for age, sex, and self-reported race. Hazard ratios per one standard deviation change in each metric are presented against adjusted p-values in volcano plots after applying Bonferroni correction for multiple comparisons.
*c. Dose–Response Analyses*: We assessed cumulative risk of incident disease across eight disease categories against varying levels of step count and sedentary time. All disease phenotypes under the same category were grouped together with equal weights given to each condition. Segmented regression was used to test for non-linear associations between levels of daily step count and sedentary time with the incidence of any phenotype within each disease category after adjusting for age, sex, and self-reported race. For categories with a statistically significant non-linear association, the elbow of the dose-response curve (inflection point) was subsequently identified.

Categorical variables are presented in frequency and percentage. Continuous variables are presented in mean and standard deviation or median and interquartile range, as appropriate. Analysis of variance (ANOVA) and Kruskal–Wallis tests were used to assess group differences, and post-hoc pairwise comparisons were performed with Bonferroni correction as appropriate. Bonferroni correction was also applied to PheWAS and dose-response analysis for multiple comparisons. All statistical tests are 2-sided with a pre-adjusted level of significance of 0.05.

Computational and statistical analyses were performed using Python 3.9^15^ and on the secure cloud-based platform of AoU Researcher Workbench^10^. All codes are available at https://github.com/broadinstitute/ml4h_aou.

## Results

In the AoU, wearable data were available for 59,018 participants, with a total of 41,721,100 person-days of monitoring over a median follow-up of 18 months. Data from 50,300 participants, including 19,845,612 person-days of monitoring over a median follow-up of 13 months, passed the quality control and were included in our final analysis (CONSORT diagrams in **Figure 1** and **Supplement, Figure S1**).

The study population was predominantly female (n= 34,258, 68%) and White (n=35,784, 71%), with a majority holding a college or advanced degree (n=31,645, 63%, **Table S1**). The median age of study participants was 55 (IQR 40-68) years, 26,723 (53%) were employed at the time of enrollment with a median income between $100k-200k. The average daily step count and sedentary time were 7,503 (95% CI 7,471-7,535) steps/day, and 591 (95% CI 590-592) min/day, respectively. Characteristics of participants with available EHRs and those in each enrollment pathway (BYOD vs WEAR) are presented in **Supplement** (**Tables S2-S3**). Adherence to wearable monitoring was higher among participants in the WEAR than those in the BYOD enrollment pathway (average wear time 1397 min/day in WEAR vs. 1393 min/day in BYOD, p-value < 0.001, **Supplement, Figure S2**). Geographic distribution of AoU participants and those with available wearable data are presented in **Supplement** (**Figure S3**). The study period included the COVID-19 pandemic, which was associated with a decrease in physical activity during the lockdown and post-lockdown periods between 2020-2022 (**Supplement, Figure S4**), similar to previous reports from wearable studies in the AoU^16^. The impact of quality control measures on the distribution of physical activity is represented in **Supplement** (**Figures S5**).

### Temporal and Geographic Patterns of Physical Activity

We assessed the patterns of step count and sedentary time across various time intervals. Minute-level step count showed a bimodal distribution with a peak around 12:00 pm and spikes at whole hour timestamps (**Figure 2A**). Aside from the expected increase in sedentary time during sleep hours, daily sedentary behavior showed a downward trend through the morning and upward trend in early afternoon with peak sedentary time at 2:57 pm.

**Figure 2.**
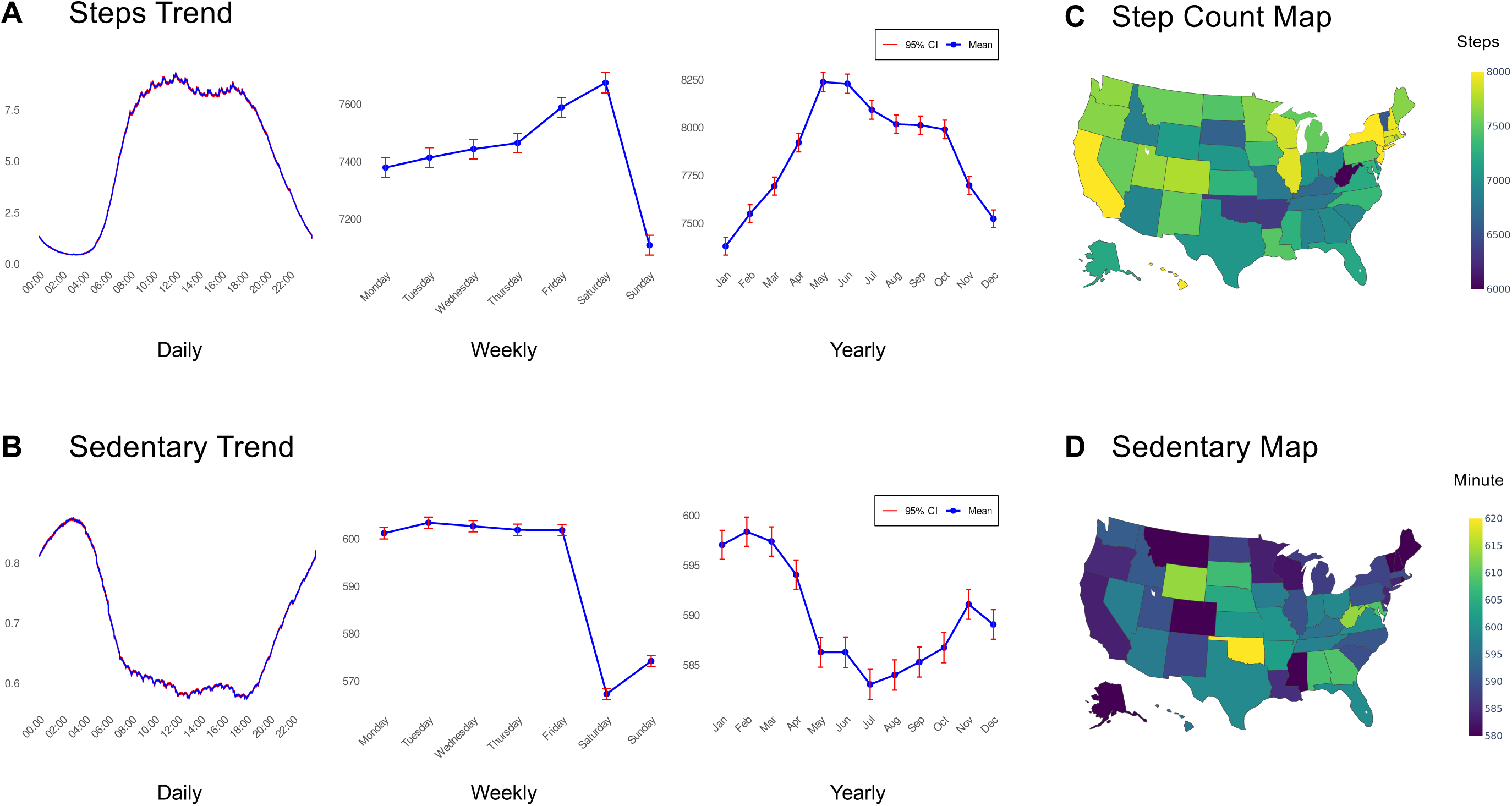
Temporal and Geographic Variations in Step Counts and Sedentary Times. (A) and (B) represent minute-level times series and aggregated distributions of step counts and sedentary times across daily, weekly, and yearly intervals. (C) and (D) illustrate geographic variations in daily average step counts and sedentary times across all U.S. states and the District of Columbia. Color scales represent the distribution of each metric across states, with yellow indicating higher and blue indicating lower values.

Participants recorded the highest daily average step count (7,674 [95%CI:7,638-7,710] steps/day) and lowest sedentary time (mean 567, 95% CI 566-568 min/day) on Saturdays, whereas Sundays exhibited the opposite pattern. Despite numerically significant differences in day-to-day variations of step count and sedentary time over different days of the month (p-value < 0.001), no consistent pattern emerged (**Supplement, Figure S6)**. However, both step count and sedentary behavior displayed significant seasonal variations (**Figure 2A-B**). Step count peaked in May (mean 8,237, 95% CI 8,187-8,287 step/day) and trended down in fall toward its lowest level in January (mean 7,379, 95% CI 7,333-7,425 per day). Conversely, sedentary time reached its highest level in February (mean 598, 95% CI 597-600 min/day), followed by a marked decline in the spring, reaching its minimum of 583 (95% CI 582-585) min/day in July. Among individuals with at least 5 years of contiguous wearable monitoring, step count trended down and sedentary time trended up in each subsequent year of follow-up (p-value < 0.001, **Supplement, Figure S6**).

We also examined geographic variations in step count and sedentary behavior across the US states (**Figure 2C and 2D**). Average daily step count was relatively higher in participants living in the Northeast, upper Midwest, and West Coast and lower in those residing in central Southern states. Step counts were highest in the District of Columbia, Hawaii, and New York, and lowest in West Virginia, Oklahoma, and Arkansas. Patterns of sedentary time showed an inverse trend with highest levels recorded in participants living in South Dakota, Kentucky, Wyoming, and Oklahoma, and lowest values observed in residents of Mississippi, Hawaii and Louisiana. Seasonal variations of step count and sedentary time across US states are presented in **Supplement** (**Figure S7**).

### Physical Activity Across Sociodemographic Subgroups

Step count demonstrated unique patterns across sociodemographic subgroups (**Figure 3**). Participants aged 18-44 and 45-65 years showed similar bimodal distributions, with peak activity in late morning and late afternoon hours. In contrast, those who were 65 years of age or older had peak step counts in the early morning with a gradual decline through the rest of the day. Males consistently recorded higher step counts than females (daily average 8,396 [95% CI 8,334-8,458] vs 7,092 [95% CI 7,056-7,129] steps/day, p-value < 0.001). Asian and White participants had higher step counts than Black participants (daily average for Asians 8,502 [95% CI 8,360-8,645], Whites 7,433 [95% CI 7,395-7,471], and Blacks 7,170 [95% CI 7,053-7,287], p-value < 0.001). In addition, Asian participants showed a unique pattern of activity with peak step count in the evening (**Figure 3**).

**Figure 3.**
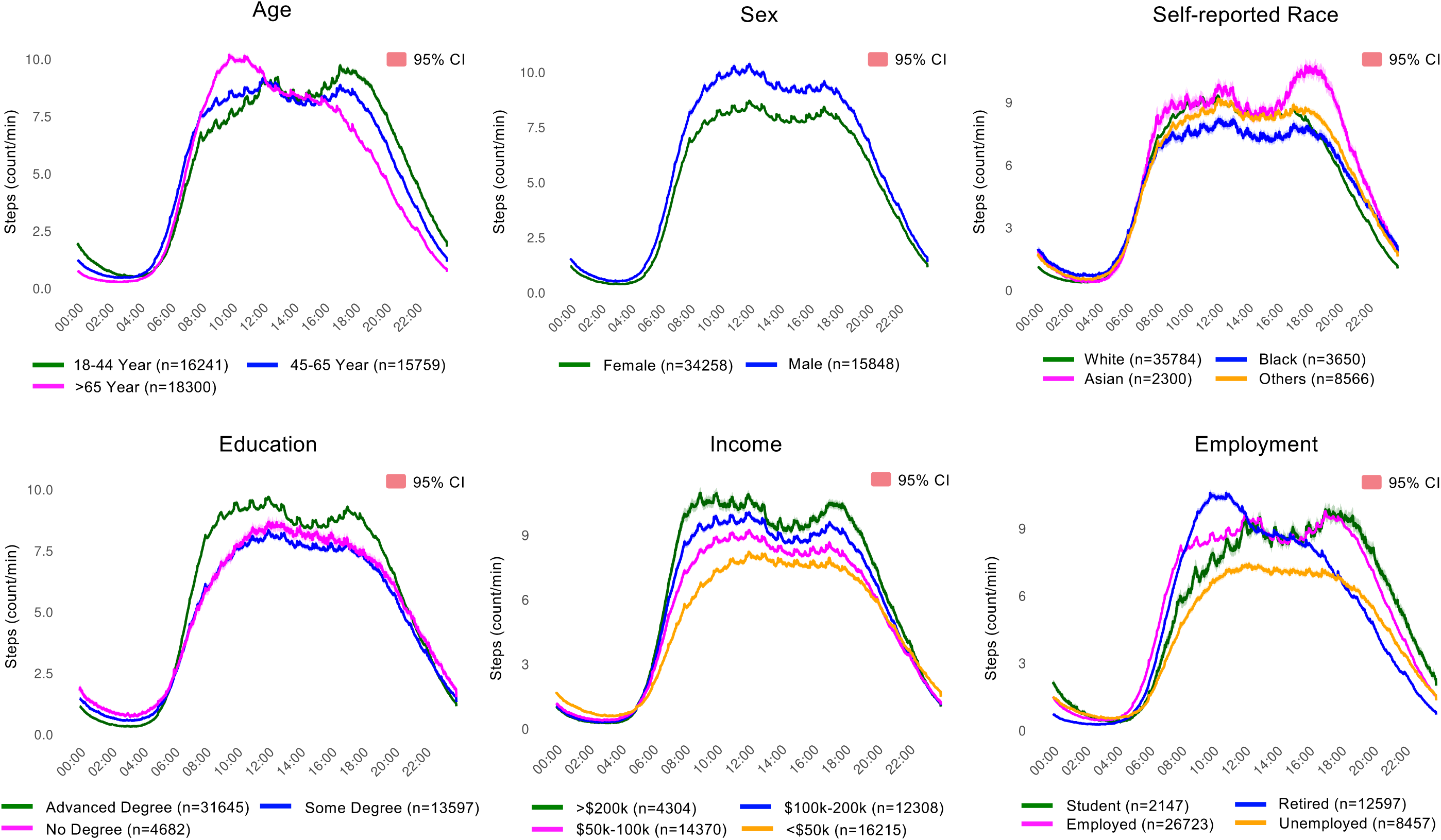
Step Counts Across Sociodemographic Subgroups. Minute-level time series of step counts shown across sociodemographic subgroups of age, sex, self-reported race, education, income, and employment status. Lines represent minute-level average step counts with 95% confidence intervals.

Step count patterns also varied by educational attainment and income. Individuals with college or advanced degrees recorded the highest step counts (mean daily step count 7,842 [95% CI 7,802-7,881] steps/day). A similar gradient was observed by income, as individuals with a household income greater than $200k had the highest step counts (mean 8,874 [95% CI 8,773-8,976] steps/day), followed by those in the $100k–200k and $50k–100k groups, with the lowest step counts observed in participants earning less than $50k. Among employment categories, retired participants had their peak step count in the early morning, while students and employed individuals showed bimodal patterns with peak step counts in the evening. Weekly patterns of step count across demographic groups are presented in **Supplement** (**Figure S8**). Temporal patterns of step count across demographic groups within BYOD and WEAR enrollment pathways are also presented in **Supplement (Figures S9-S12)**.

### PheWAS and Dose-Response Analysis

We performed a PheWAS analysis to assess the relationship between step count and sedentary time with incident clinical phenotypes across eight disease categories (**Figure 4**). In this prospective analysis, the median follow up time was 16 months for incident cases and 29 months for controls. Higher step counts were significantly associated with lower incidence of multiple conditions across the disease spectrum, including obesity (HR = 0.60 [95% CI 0.56-0.64]), sleep apnea (HR = 0.64 [0.60-0.68]), abnormalities of breathing (HR = 0.72 [0.68-0.77]), hypotension (HR = 0.49 [0.43-0.56]), gastroesophageal reflux (HR = 0.72 [0.67-0.76]), mood disorders (HR = 0.72 [0.67-0.77]) and myalgia (HR = 0.71 [0.65-0.77]). In contrast, longer sedentary times were associated with higher incidence of morbid obesity (HR = 1.85 [1.72-2.00]), sleep apnea (HR = 1.41 [1.33-1.50]), essential hypertension (HR = 1.27 [1.21-1.34]), hypoxemia (HR = 1.59 [1.38-1.83]), fatty liver disease (HR = 1.31 [1.20-1.44]) and dysthymic disorders (HR = 1.46 [1.25-1.70]). Detailed results of our phenome-wide association analysis are presented in **Supplement** (**Tables S4-S5**).

**Figure 4.**
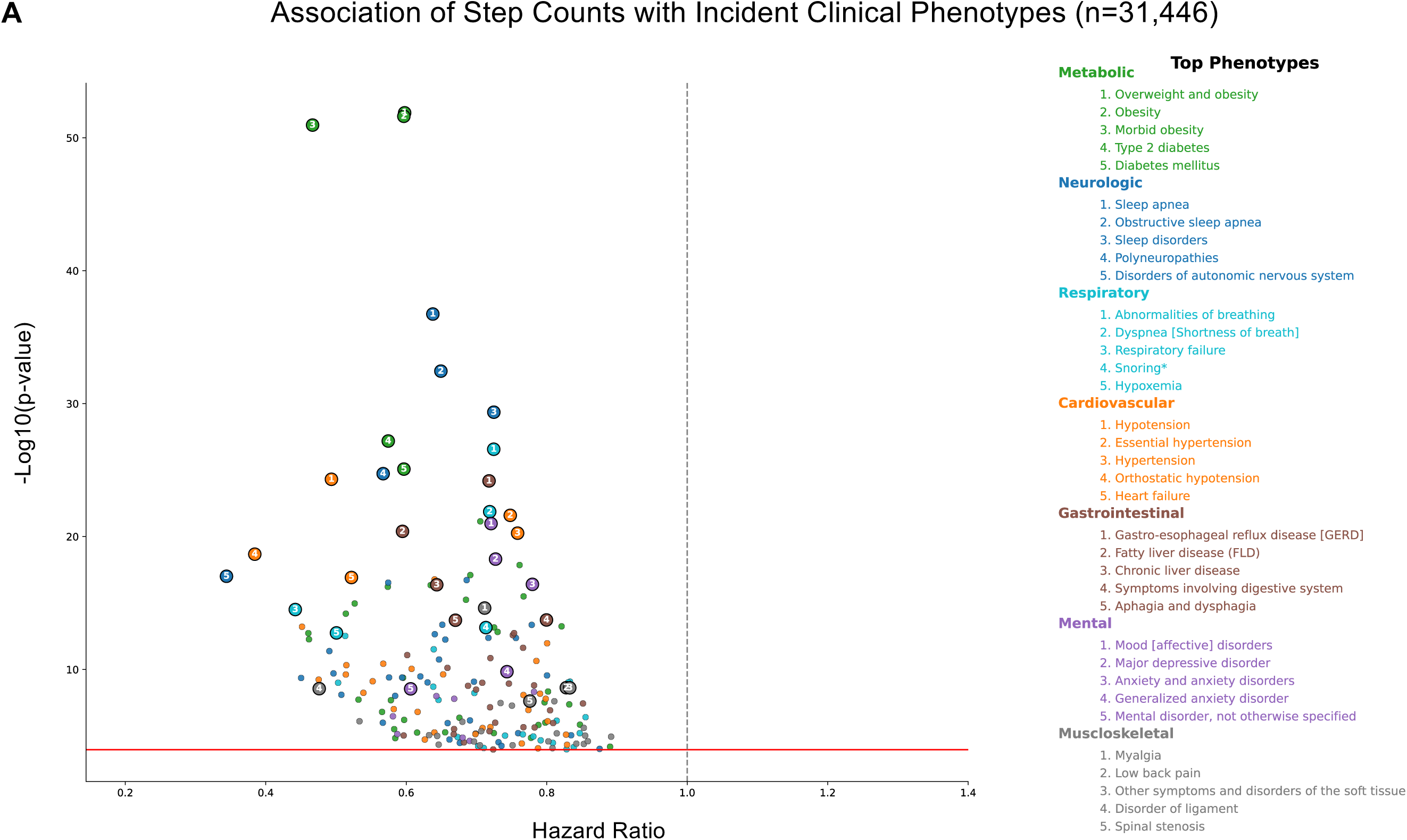
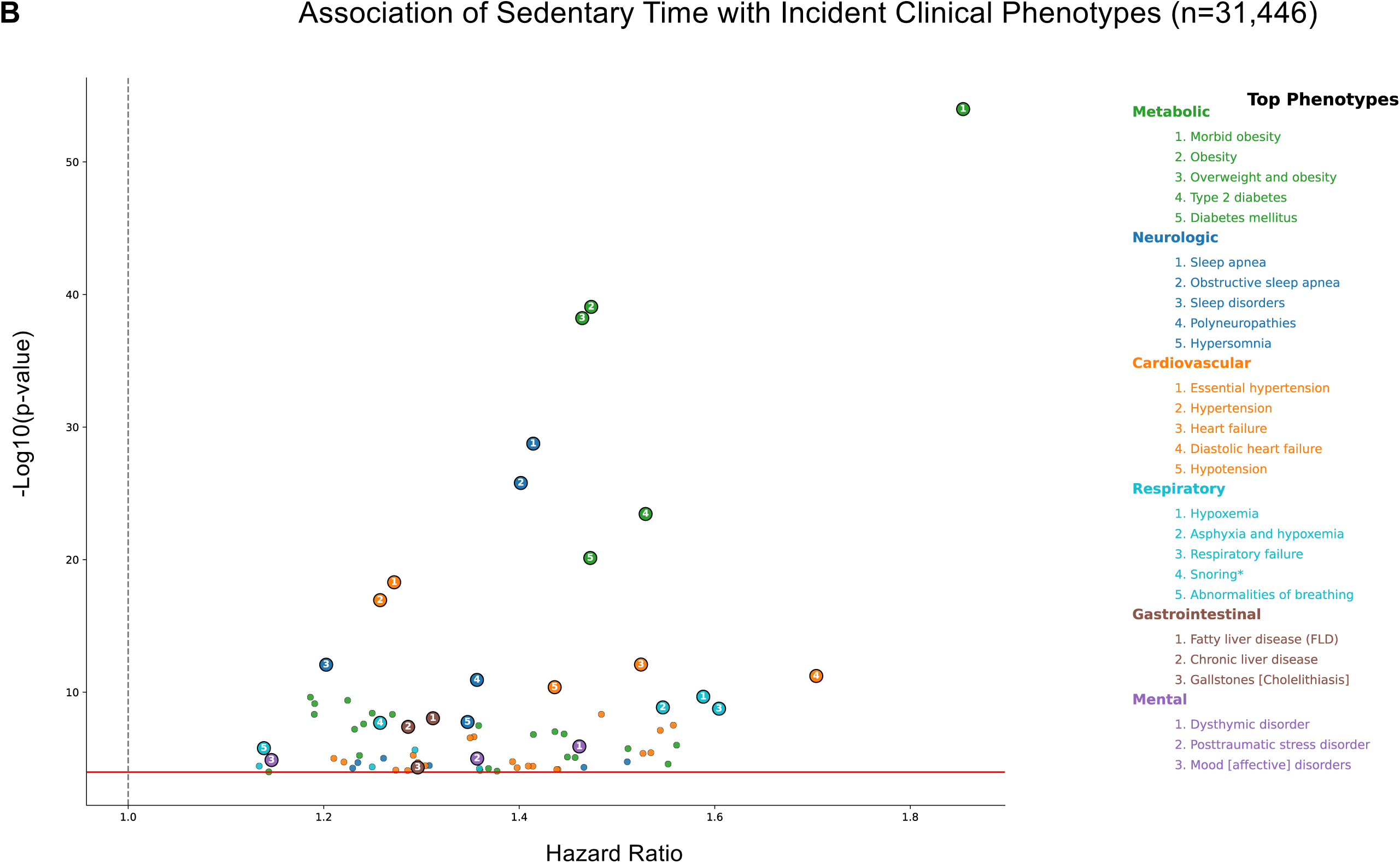
Association of Step Count and Sedentary Time with Incident Disease. Associations between daily step counts (A) and sedentary times (B) with incident phenotypes are shown across eight major disease categories. Each point represents an individual phenotype, plotted by the hazard ratio (x-axis, per one standard deviation change in each metric) and the corresponding –log10(p-values) on the y-axis. Vertical lines denote hazard ratio of 1.0 and horizontal lines indicate the Bonferroni-corrected significance threshold. All models are adjusted for age, sex, and self-reported race. Phenotypes are grouped and color-coded by disease category and the top 5 phenotypes by significance are listed where available. Among 31,661 participants with wearable data and electronic health records, the last clinical encounter or demographic covariates could not be adjudicated in 215 participants who were excluded from this analysis (n=31,446).

To better understand the nature of the relationships between physical activity and sedentary behavior with clinical outcomes, we performed a dose-response analysis between daily step count and sedentary time with cumulative risk of incident disease across eight disease groups. Increasing numbers of daily steps and shorter duration of sedentary time were associated with lower incidence of all eight disease categories (p for trend < 0.001 for all, **Figure 5**). The inverse association between step count and disease groups plateaued beyond approximately 11,000 to 12,000 steps/day for neurologic disorders (p-value <0.001) and 9,000 to 10,000 steps/day for cardiovascular diseases (p-value<0.001). Similarly, the association between daily sedentary time and neurologic, cardiovascular, and metabolic diseases reached a plateau below 600 to 640 min/day of sedentary behavior. For all other disease groups, we did not find a statistically significant risk threshold for either step count or sedentary time (**Supplement Table S6-S7**). Dose-response associations between deciles of these two metrics and incident disease categories are presented in **Supplement** (**Figure S13**).

**Figure 5.**
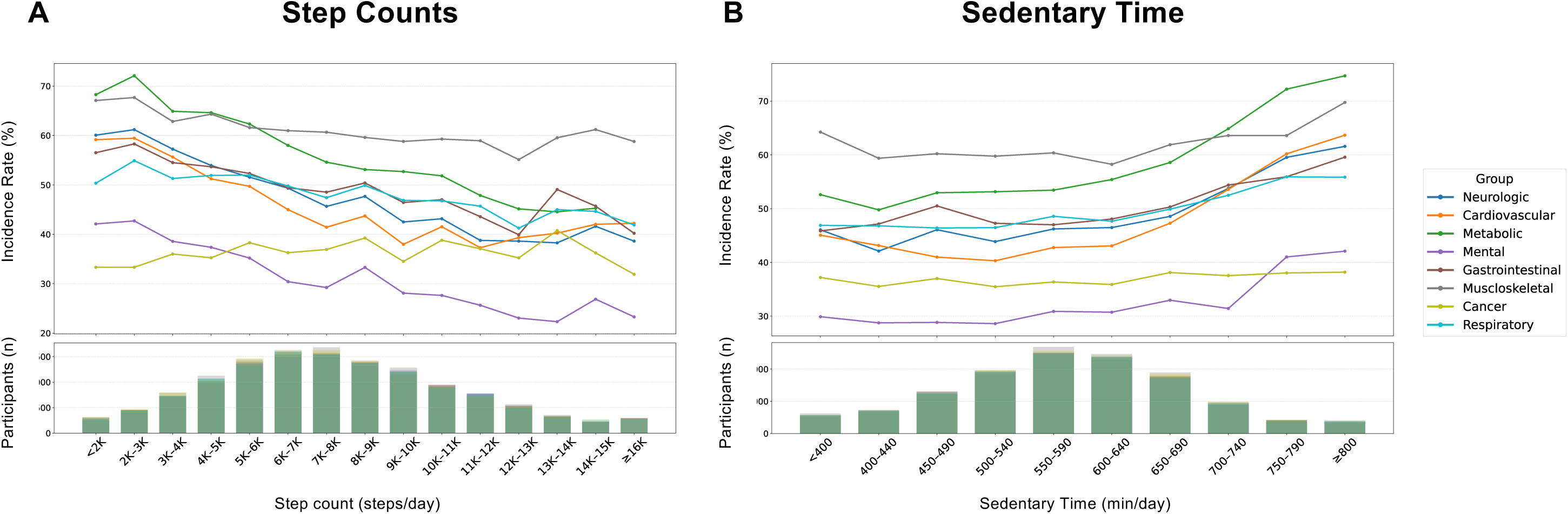
Dose-Response Analysis of Daily Step Counts and Sedentary Time. The figures show cumulative incidence rate of eight major disease categories across varying levels of daily step counts (A) and sedentary time (B). Histograms at the bottom of each panel indicate the number of participants contributing data to each exposure bin.

## Discussion

Leveraging data from over 19 million person-days of wearable monitoring in broad populations, this study represents one of the largest longitudinal analyses of physical activity with minute-level resolution and multi-year follow-up. Our findings provide a detailed picture of temporal and geographic patterns of physical activity and sedentary behavior among US adults, including their variations across sociodemographic groups. Our PheWAS analysis complements prior studies on the association of step count and sedentary behavior with clinical outcomes. Moreover, we identify daily thresholds for step count and sedentary time that are associated with the lowest observed risk of disease across the spectrum of clinical conditions. Collectively, these findings provide a comprehensive picture of physical activity and sedentary behavior across the US and can potentially inform actionable recommendations in future guidelines to improve population health.

We observed substantial temporal variations in both physical activity and sedentary behavior. Step count generally followed a bimodal daily distribution, peaking at late morning, followed by lower activities in mid-afternoon hours before reaching a second peak in the evening. We also observed higher levels of physical activity on Saturdays and a seasonal trend with greater activity during spring and summer. Temporal patterns of physical activity are known to correlate with distinct risk profiles of obesity and metabolic syndrome^17^, highlighting the importance of characterizing these patterns at the population level. Our observations are also consistent with previous studies on environmental and behavioral factors, such as daylight availability, work schedules, and weather conditions, which can significantly influence the patterns of physical activity^18^.

We also observed substantial variations in physical activity across demographic subgroups which highlight the influence of structural and societal factors on health-related behaviors. Our findings support prior reports on the association of demographic features with physical activity in other populations^19,20^, while offering a comprehensive characterization of such patterns among US adults. Geographic variations of physical activity similarly reflect a complex interaction of individual characteristics, built environments, and broader societal factors that shape opportunities for physical activity across the US. Notably, we observed differential seasonal patterns between northern and southern states, highlighting the significant role of environmental factors on physical activity. The breadth of data collected in the AoU, including longitudinal surveys of social determinants of health and linkage to geospatial environmental exposures, empowers future research to investigate the multifaceted interactions between individual characteristics, environmental conditions, and health behaviors at scale.

The significance of these observations is underscored by the association of physical activity and sedentary behavior with a broad range of clinical outcomes. In our analysis, cardiometabolic disorders such as obesity and hypertension, and psychiatric diseases such as mood and anxiety disorders consistently stood out as correlates of inactivity and sedentary behavior. Consistent associations between step count and sedentary time with incident disease, together with findings from our dose-response analyses reinforce the existing literature that associates physical inactivity with poor health outcomes^21–24^. Furthermore, our analysis demonstrates the heterogeneous relationship of varying levels of step counts and sedentary time with clinical outcomes. In many disease groups, these relationships appear to be linear, with no limit to the benefits of higher step counts and lower sedentary times. In contrast, the cardiovascular and neurologic benefits appear to plateau beyond certain thresholds of daily step counts and sedentary times. Across disease categories, graded associations between physical activity metrics and incident disease revealed distinct nonlinear patterns, enabling identification of step count and sedentary time thresholds associated with lower disease risk. These findings suggest disease-specific relationships between step counts and sedentary time with clinical outcomes, with both linear and threshold-dependent patterns that can inform personalized guidance for different populations in future.

At a broader scale, our study demonstrates the value of wearables to characterize dynamic patterns of physical activity in real-world settings^25^. The richness of wearable data extends beyond self-reported estimates of physical activity or aggregate measures of daily exercise. Importantly, current guidelines do not universally recognize wearable technology as a tool for measurement and promotion of physical activity^2,3^. Wearable monitoring can inform a data-driven approach to understand behavior-disease relationships when linked with EHRs in large-scale cohorts such as AoU^26^. Although integration of wearable data into clinical workflows may not be straightforward^27^, a wearable-centered approach has the potential to facilitate the design of tailored interventions for promotion of physical activity in target populations. Because step count and sedentary time are routinely captured by consumer wearables, these metrics offer intuitive and monitorable targets that may complement existing physical activity counseling in clinical and public health settings.

### Limitations

First, the study sampling is prone to selection bias as it is more likely for health-conscious individuals who use wearables to be enrolled through the BYOD pathway. The AoU attempts to mitigate this bias by enrolling a community-based cohort through the WEAR study. However, the selection process remains non-random, and the study sample is not representative of the broader US population^28^. Second, characterization of clinical phenotypes relies on EHRs which are inherently subject to incomplete and variable adjudication at local sites, particularly as data are primarily collected for clinical and not research indications. Third, participating health systems follow varying measurement and documentation standards, introducing heterogeneity in clinical phenotypes that may affect the consistency of covariate or outcome ascertainment. Fourth, wearable data in this study were derived exclusively from Fitbit devices; therefore, the observed patterns and associations may not generalize to users of other vendors with different sensors or algorithms. Finally, this study is observational in nature and despite our attempts for prospective investigation in PheWAS and dose-response analysis, causal inferences between patterns of physical activity and clinical outcomes cannot be definitively established. Despite these limitations, the large sample size, longitudinal follow-up, and high-resolution wearable data provide robust evidence of associations between physical activity and incident disease.

### Conclusions

This study provides a large-scale, high-resolution characterization of physical activity and sedentary behavior among U.S. adults in the All of Us Program. By analyzing more than 19 million person-days of wearable monitoring, we identified temporal, geographic, and sociodemographic patterns of physical activity and demonstrated associations with incident disease across multiple clinical domains, including disease-specific nonlinear relationships. In particular, cardiovascular risk reduction plateaued at approximately 9,000 to 10,000 steps per day, while higher disease risk was observed beyond approximately 600 to 640 minutes per day of sedentary time. These findings underscore the value of integrating granular wearable data with electronic health records to inform more actionable physical activity guidance and population-level disease prevention strategies.

## Data Availability

All data area available online at the Research Workbench from the All of Us Research Program

## Acknowledgment

We gratefully acknowledge *All of Us* participants for their contributions, without whom this research would not have been possible. We also thank the National Institutes of Health’s *All of Us* program for making available the participant data used in this study.

