## Supplements for "Patterns and Clinical Outcomes of Physical Activity and Sedentary Behavior Across 20 Million Days of Wearable Monitoring in U.S. Adults"

### Supplemental Material

Index

Method Description: Quality control
Method Description: Definition of sedentary activity
Table S1. Baseline Characteristics of Study Population.
Table S2. Baseline characteristics of participants with available electronic health records (EHRs).
Table S3. Baseline characteristics of enrolled participants in the BYOD and WEAR pathways.
Table S4. Phenome-wide association analysis of step counts
Table S5. Phenome-wide association analysis of sedentary time
Table S6. Non-linearity and threshold analysis of step count and disease incidence
Table S7. Non-linearity and threshold analysis of sedentary time and disease incidence
Figure S1. CONSORT flow diagram of enrollment pathways
Figure S2. Adherence to wearable monitoring across enrollment pathways
Figure S3. Geographic distribution of All of Us Research Program participants
Figure S4. Impact of COVID-19 pandemic on step count
Figure S5. Impact of quality control on step count distribution
Figure S6. Monthly and multi-year variations of step count and sedentary time
Figure S7. Seasonal Variations of Step Count and Sedentary Time Across US States.
Figure S8. Weekly step counts across sociodemographic subgroups
Figure S9. Daily step counts in the BYOD enrollment pathway
Figure S10. Daily step counts in the WEAR enrollment pathway
Figure S11. Weekly step counts in the BYOD enrollment pathway
Figure S12. Weekly step counts in the WEAR enrollment pathway
Figure S13. Dose-Response analysis across deciles of daily step count and sedentary time.

**Methods Description: Quality Control**

In this analysis, we used minute-level step counts and heart rates to assess the patterns of physical activity and sedentary behavior. The raw minute-level data are noisy and require substantial preprocessing to ensure data quality for downstream analysis:

1- Data vectorization: Minute-level step count and heart rate measurements of each participant were transformed into arrays of shape [1440, 1] from midnight to midnight.

2- Non-wear: Periods with no corresponding heart rates were considered as non-wear times and excluded from analysis of step counts.

3- Outliers: Step count values of greater than 250 steps/minute and heart rate values outside the range of 30-200 beats/minute were excluded from analysis. These thresholds were derived from visual inspection of data distributions to identify plausible physiological ranges for step count and heart rate.

4- Missingness: Daily arrays containing more than 200 missing data points, equivalent to less than 3 hours and 20 minutes of data collection per day, were excluded. This threshold corresponded to the 1$^{st}$ percentile of step count and heart rate data distributions.

Notably, heart rate data from 11,849 participants, including a total of 3,713,640 person-days of monitoring were reported at second-level or fractions of minutes. This included heart rates from 11,034 participants, including 2,541,888 person-days of monitoring which passed our quality control and were used in the final analysis. All sub-minute data were converted to minute-level averages to maintain consistency of downstream analysis.

**Methods Description: Sedentary Activity**

Clinical guidelines of physical activity define sedentary behavior as activities with metabolic equivalents (METs) of 1.5 or less but do not provide a non-metabolic definition applicable to wearables[1]. In addition, Fitbit trackers only provide daily aggregates of sedentary times. To quantify sedentary behavior at minute-level resolution, we identified sedentary times using accelerometry and heart rate data. These calculations were based on estimates of maximum heart rate ($HR_{max}$), resting heart rate ($HR_{rest}$), and heart rate reserve (HRR). Sleep time was removed from this analysis. Specifically, sedentary time was defined using a combined criterion:

(1) minute-level step count of zero, and

(2) concurrent HRR of less than 20%.

This hybrid definition integrates both movement and physiological signals to more accurately capture sedentary behavior from wearable devices.

$HR_{max}$ was estimated using the following formula:

$$HR_{max} = 220 - \text{age}$$

In subsequent years of follow up, the calculated $HR_{max}$ was adjusted for chronological increase in participant age. $HR_{rest}$ was estimated from the mode value of minute-level heart rate readings. HRR was then calculated as the difference between the maximum and resting heart rates:

$$HRR = HR_{max} – HR_{rest}$$

Notably, earlier generations of Fitbit devices use proprietary algorithms that estimate physical activity zones from $HR_{max}$ and not HRR[2]. To ensure consistency, we applied the HRR-based approach to data from all participants regardless of the device version. This definition is intended to capture *relative sedentary behavior* rather than absolute MET-based inactivity.

**Table S1. Baseline Characteristics of Study Population.** Data are presented in numbers (percentages). For clinical characteristics, proportions are calculated from a denominator who had available electronic health records (n=31,661).

| | Study Population (n=50,300) |
|---|---|
| Age | |
| 18-44 (years) | 16,241 (32.3%) |
| 45-65 (years) | 18,300 (36.4%) |
| >65 (years) | 15,759 (31.3%) |
| Sex Assigned at Birth | |
| Female | 34,258 (68.1%) |
| Male | 15,848 (31.5%) |
| Race | |
| White | 35,784 (71.1%) |
| Black or African American | 3,650 (7.3%) |
| Asian | 2,300 (4.6%) |
| Others | 8,566 (17.0%) |
| Ethnicity | |
| Hispanic or Latino | 5,653 (11.2%) |
| Non-Hispanic or Latino | 43,611 (86.7%) |
| Others | 1,036 (2.1%) |
| Education | |
| College or Advanced Degree | 31,645 (62.9%) |
| Some Degree | 13,597 (27.0%) |
| No Degree | 4,682 (9.3%) |
| Others | 376 (0.7%) |
| Smoking | |
| Never | 30,660 (61.0%) |
| Ever (>=100 cigarettes) | 17,968 (35.7%) |
| Other | 1,672 (3.3%) |
| Alcohol | |
| Never | 7,511 (14.9%) |
| Monthly or less | 16,111 (32.1%) |
| 2-4 times/month | 10,286 (20.4%) |
| 2-3 times/week | 6,916 (13.7%) |
| >= 4 times/week | 6,448 (12.8%) |
| Others | 3,028 (6.1%) |
| Body Mass Index (BMI) | |
| BMI <= 25 | 12,633 (29.1%) |
| BMI 25 - 30 | 13,244 (30.5%) |
| BMI > 30 | 17,573 (40.4%) |
| Hypertension | 11,589 (36.6%) |
| Diabetes | 4,802 (15.2%) |
| Hyperlipidemia or Dyslipidemia | 11,745 (37.1%) |
| Coronary Artery Disease | 2,647 (8.4%) |
| Heart Failure | 1,521 (4.8%) |
| Chronic Kidney Disease | 2,115 (6.7%) |

**Table S2. Baseline Characteristics of Participants with Available Electronic Health Records (EHRs).** Data are presented in numbers (percentages).

| | Participants with EHRs (n=31,661) |
|---|---|
| **Age** | |
| **18-44 (years)** | 9,408 (29.7%) |
| **45-65 (years)** | 11,442 (36.2%) |
| **>65 (years)** | 10,811 (34.1%) |
| **Sex Assigned at Birth** | |
| **Female** | 21,510 (67.9%) |
| **Male** | 10,022 (31.7%) |
| **Race** | |
| **White** | 22,616 (71.4%) |
| **Black or African American** | 2,467 (7.8%) |
| **Asian** | 1,370 (4.4%) |
| **Others** | 5,208 (16.4%) |
| **Ethnicity** | |
| **Hispanic or Latino** | 3,374 (10.7%) |
| **Non-Hispanic or Latino** | 27,615 (87.2%) |
| **Others** | 672 (2.1%) |
| **Education** | |
| **College or Advanced Degree** | 19,761 (62.4%) |
| **Some Degree** | 8,736 (27.6%) |
| **No Degree** | 2,954 (9.3%) |
| **Others** | 210 (0.7%) |
| **Smoking** | |
| **Never** | 19,439 (61.4%) |
| **Ever (>=100 cigarettes)** | 11,613 (36.7%) |
| **Other** | 609 (1.9%) |
| **Alcohol** | |
| **Never** | 4,830 (15.3%) |
| **Monthly or less** | 10,152 (32.1%) |
| **2-4 times per month** | 6,661 (21.0%) |
| **2-3 times per week** | 4,472 (14.1%) |
| **>= 4 times per week** | 4,174 (13.2%) |
| **Others** | 1,372 (4.3%) |
| **Body Mass Index (BMI)** | |
| **BMI <= 25** | 8,157 (27.5%) |
| **BMI 25 - 30** | 9,097 (30.7%) |
| **BMI > 30** | 12,364 (41.8%) |
| **Hypertension** | 11,589 (36.6%) |
| **Diabetes** | 4,802 (15.2%) |
| **Hyperlipidemia or Dyslipidemia** | 11,745 (37.1%) |
| **Coronary Artery Disease** | 2,647 (8.4%) |
| **Heart Failure** | 1,521 (4.8%) |
| **Chronic Kidney Disease** | 2,115 (6.7%) |

**Table S3. Baseline Characteristics of Enrolled Participants in the BYOD and WEAR Pathways.** Data are presented in numbers (percentages). For clinical characteristics, proportions are calculated from a denominator who had available electronic health records (n=14,411 in BYOD and n=17,250 in WEAR).

| | BYOD (n=24,925) | WEAR (n=25,375) | p-value |
|---|---|---|---|
| **Age** | | | <0.001 |
| **18-44 (years)** | 8,005 (32.1%) | 8,236 (32.5%) | |
| **45-65 (years)** | 10,090 (40.5%) | 8,210 (32.4%) | |
| **>65 (years)** | 6,830 (27.4%) | 8,929 (35.1%) | |
| **Sex Assigned at Birth** | | | <0.001 |
| **Female** | 17,583 (70.5%) | 16,675 (65.7%) | |
| **Male** | 7,258 (29.1%) | 8,590 (33.9%) | |
| **Race** | | | <0.001 |
| **White** | 20,571 (82.5%) | 15,213 (60.0%) | |
| **Black or African American** | 1,181 (4.8%) | 2,469 (9.7%) | |
| **Asian** | 679 (2.7%) | 1,621 (6.4%) | |
| **Others** | 2,494 (10.0%) | 6,072 (23.9%) | |
| **Ethnicity** | | | <0.001 |
| **Hispanic or Latino** | 1,578 (6.3%) | 4,075 (16.1%) | |
| **Non-Hispanic or Latino** | 22,972 (92.2%) | 20,639 (81.3%) | |
| **Others** | 375 (1.5%) | 661 (2.6%) | |
| **Education** | | | <0.001 |
| **College or Advanced Degree** | 17,260 (69.2%) | 14,385 (56.7%) | |
| **Some Degree** | 5,824 (23.4%) | 7,773 (30.6%) | |
| **No Degree** | 1,638 (6.6%) | 3,044 (12.0%) | |
| **Others** | 203 (0.8%) | 173 (0.7%) | |
| **Smoking** | | | <0.001 |
| **Never** | 15,639 (62.7%) | 15,021 (59.2%) | |
| **Ever (>=100 cigarettes)** | 8,049 (32.3%) | 9,919 (39.1%) | |
| **Other** | 1,237 (5.0%) | 435 (1.7%) | |
| **Alcohol** | | | <0.001 |
| **Never** | 3,025 (12.1%) | 4,486 (17.7%) | |
| **Monthly or less** | 7,280 (29.2%) | 8,831 (34.8%) | |
| **2-4 times per month** | 5,524 (22.2%) | 4,762 (18.8%) | |
| **2-3 times per week** | 3,952 (15.9%) | 2,964 (11.7%) | |
| **>= 4 times per week** | 3,440 (13.8%) | 3,008 (11.8%) | |
| **Others** | 1,704 (6.8%) | 1,324 (5.2%) | |
| **Body Mass Index (BMI)** | | | <0.001 |
| **BMI <= 25** | 5,788 (28.9%) | 6,845 (29.2%) | |
| **BMI 25 - 30** | 6,371 (31.8%) | 6,873 (29.4%) | |
| **BMI > 30** | 7,881 (39.3%) | 9,692 (41.4%) | |
| **Hypertension** | 4,947 (34.3%) | 6,642 (38.5%) | <0.001 |
| **Diabetes** | 1,886 (13.1%) | 2,916 (16.9%) | <0.001 |
| **Hyperlipidemia or Dyslipidemia** | 5,220 (36.2%) | 6,525 (37.8%) | <0.001 |
| **Coronary Artery Disease** | 1,103 (7.7%) | 1,544 (9.0%) | <0.001 |
| **Heart Failure** | 594 (4.1%) | 927 (5.4%) | <0.001 |
| **Chronic Kidney Disease** | 787 (5.5%) | 1,328 (7.7%) | <0.001 |

**Table S4. Phenome-wide Association Analysis of Step Counts.** Associations of average daily step count with top 5 clinical phenotypes by significance in each disease category are presented. Associations are derived from Cox regression models after adjustment for age, sex, and self-reported race. Hazard ratios represent the change in each disease incidence risk for one standard deviation increase in step count. Abbreviations: HR, hazard ratio; CI, confidence interval.

| Disease Group | Phenotype | HR [95% CI] | p-value |
|---|---|---|---|
| Endocrine-Metabolic | **Overweight and obesity** | 0.6 [0.56-0.64] | 1.31e-52 |
| | **Obesity** | 0.6 [0.56-0.64] | 2.39e-52 |
| | **Morbid obesity** | 0.47 [0.42-0.52] | 1.10e-51 |
| | **Type 2 diabetes** | 0.57 [0.52-0.63] | 6.45e-28 |
| | **Diabetes mellitus** | 0.6 [0.54-0.66] | 8.34e-26 |
| Neurologic | **Sleep apnea** | 0.64 [0.6-0.68] | 1.80e-37 |
| | **Obstructive sleep apnea** | 0.65 [0.61-0.7] | 3.56e-33 |
| | **Sleep disorders** | 0.72 [0.69-0.77] | 4.36e-30 |
| | **Polyneuropathies** | 0.57 [0.51-0.63] | 1.85e-25 |
| | **Disorders of autonomic nervous system** | 0.34 [0.27-0.44] | 9.66e-18 |
| Respiratory | **Abnormalities of breathing** | 0.72 [0.68-0.77] | 2.68e-27 |
| | **Dyspnea [Shortness of breath]** | 0.72 [0.67-0.77] | 1.34e-22 |
| | **Respiratory failure** | 0.44 [0.36-0.54] | 3.07e-15 |
| | **Snoring*** | 0.71 [0.65-0.78] | 7.02e-14 |
| | **Hypoxemia** | 0.5 [0.42-0.6] | 1.77e-13 |
| Cardiovascular | **Hypotension** | 0.49 [0.43-0.56] | 4.87e-25 |
| | **Essential hypertension** | 0.75 [0.71-0.79] | 2.51e-22 |
| | **Hypertension** | 0.76 [0.72-0.8] | 5.51e-21 |
| | **Orthostatic hypotension** | 0.38 [0.31-0.47] | 2.10e-19 |
| | **Heart failure** | 0.52 [0.45-0.61] | 1.20e-17 |
| Gastrointestinal | **Gastro-esophageal reflux disease [GERD]** | 0.72 [0.67-0.76] | 6.46e-25 |

| | | | |
|---|---|---|---|
| | **Fatty liver disease (FLD)** | 0.59 [0.53-0.66] | 4.04e-21 |
| | **Chronic liver disease** | 0.64 [0.58-0.71] | 4.20e-17 |
| | **Symptoms involving digestive system** | 0.8 [0.76-0.85] | 1.87e-14 |
| | **Aphagia and dysphagia** | 0.67 [0.6-0.74] | 1.96e-14 |
| **Psychiatric** | **Mood [affective] disorders** | 0.72 [0.67-0.77] | 1.05e-21 |
| | **Major depressive disorder** | 0.73 [0.68-0.78] | 4.93e-19 |
| | **Anxiety and anxiety disorders** | 0.78 [0.74-0.83] | 3.92e-17 |
| | **Generalized anxiety disorder** | 0.74 [0.68-0.81] | 1.46e-10 |
| | **Mental disorder, not otherwise specified** | 0.61 [0.51-0.72] | 2.86e-09 |
| **Musculoskeletal** | **Myalgia** | 0.71 [0.65-0.77] | 2.38e-15 |
| | **Low back pain** | 0.83 [0.78-0.88] | 2.34e-09 |
| | **Other symptoms and disorders of the soft tissue** | 0.83 [0.78-0.88] | 2.38e-09 |
| | **Disorder of ligament** | 0.48 [0.37-0.61] | 2.77e-09 |
| | **Spinal stenosis** | 0.78 [0.71-0.85] | 2.38e-08 |

**Table S5. Phenome-wide Association Analysis of Sedentary Time.** Associations of average daily sedentary time with top 5 clinical phenotypes by significance in each disease category are presented. For disease categories where fewer than 5 associations were significant, all significant associations are presented. Associations are derived from cox regression models after adjustment for age, sex, and self-reported race. Hazard ratios represent the change in each disease incidence risk for one standard deviation increase in sedentary time. Abbreviations: HR, hazard ratio.

| Disease Group | Phenotype | HR [95% CI] | p-value |
|---|---|---|---|
| Endocrine-Metabolic | Morbid obesity | 1.85 [1.72-2.0] | 1.04e-54 |
| | Obesity | 1.47 [1.39-1.56] | 8.60e-40 |
| | Overweight and obesity | 1.46 [1.38-1.55] | 6.05e-39 |
| | Type 2 diabetes | 1.53 [1.41-1.66] | 3.61e-24 |
| | Diabetes mellitus | 1.47 [1.36-1.6] | 7.53e-21 |
| Neurologic | Sleep apnea | 1.41 [1.33-1.5] | 1.78e-29 |
| | Obstructive sleep apnea | 1.4 [1.32-1.49] | 1.65e-26 |
| | Sleep disorders | 1.2 [1.14-1.26] | 8.45e-13 |
| | Polyneuropathies | 1.36 [1.24-1.48] | 1.18e-11 |
| | Hypersomnia | 1.35 [1.21-1.49] | 1.77e-08 |
| Cardiovascular | Essential hypertension | 1.27 [1.21-1.34] | 5.13e-19 |
| | Hypertension | 1.26 [1.19-1.33] | 1.14e-17 |
| | Heart failure | 1.52 [1.36-1.71] | 8.20e-13 |
| | Diastolic heart failure | 1.7 [1.46-1.98] | 5.98e-12 |
| | Hypotension | 1.44 [1.29-1.6] | 4.21e-11 |
| Respiratory | Hypoxemia | 1.59 [1.38-1.83] | 2.20e-10 |
| | Asphyxia and hypoxemia | 1.55 [1.34-1.78] | 1.41e-09 |
| | Respiratory failure | 1.6 [1.38-1.87] | 1.76e-09 |
| | Snoring* | 1.26 [1.16-1.36] | 2.05e-08 |
| | Abnormalities of breathing | 1.14 [1.08-1.2] | 1.67e-06 |

| | | | |
|---|---|---|---|
| **Gastrointestinal** | **Fatty liver disease (FLD)** | 1.31 [1.2-1.44] | 9.26e-09 |
| | **Chronic liver disease** | 1.29 [1.18-1.41] | 4.11e-08 |
| | **Gallstones [Cholelithiasis]** | 1.3 [1.14-1.47] | 4.75e-05 |
| **Psychiatric** | **Dysthymic disorder** | 1.46 [1.25-1.7] | 1.24e-06 |
| | **Posttraumatic stress disorder** | 1.36 [1.18-1.55] | 1.01e-05 |
| | **Mood [affective] disorders** | 1.15 [1.08-1.22] | 1.32e-05 |

**Table S6. Non-linearity and Threshold Analysis of Step Count and Disease Incidence.** The table represents the results of non-linearity and threshold analysis of the associations between average daily step counts and cumulative risk of incident disease across eight categories of clinical phenotypes. All disease phenotypes under the same category were grouped together with equal weights for this cumulative analysis. Segmented regression was used to test for a non-linear dose-response association between daily step counts and the incidence of any phenotype within each disease category (p-value). All models were adjusted for age, sex, and self-reported race. Bonferroni correction was applied to the significance threshold to account for multiple comparisons in both step count and sedentary time analysis (adjusted alpha: 0.0031). For categories with a significant non-linear association, the elbow of the dose-response curve (inflection point) was subsequently identified.

| Disease Category | Non-Linear Association | p-value | Inflection Point |
|---|---|---|---|
| **Neurologic** | Significant | <0.001 | 11k–12k |
| **Cardiovascular** | Significant | <0.001 | 9k–10k |
| **Metabolic** | Non-significant | 0.037 | - |
| **Mental** | Non-significant | 0.026 | - |
| **Gastrointestinal** | Non-significant | 0.2 | - |
| **Musculoskeletal** | Non-significant | 0.014 | - |
| **Cancer** | Non-significant | 0.006 | - |
| **Respiratory** | Non-significant | 0.022 | - |

**Table S7. Non-linearity and Threshold Analysis of Sedentary Time and Disease Incidence.** The table represents the results of non-linearity and threshold analysis of the associations between average daily sedentary time and cumulative risk of incident disease across eight categories of clinical phenotypes. All disease phenotypes under the same category were grouped together with equal weights for this cumulative analysis. Segmented regression was used to test for a non-linear dose-response association between daily sedentary time and the incidence of any phenotype within each disease category (p-value). All models were adjusted for age, sex, and self-reported race. Bonferroni correction was applied to the significance threshold to account for multiple comparisons in both step count and sedentary time (adjusted alpha: 0.0031). For categories with a significant non-linear association, the elbow of the dose-response curve (inflection point) was subsequently identified.

| Disease Category | Non-Linear Association | p-value | Inflection Point |
|---|---|---|---|
| **Neurologic** | Significant | <0.001 | 600–640 |
| **Cardiovascular** | Significant | <0.001 | 600–640 |
| **Metabolic** | Significant | <0.001 | 600–640 |
| **Mental** | Non-significant | 0.006 | - |
| **Gastrointestinal** | Non-significant | 0.004 | - |
| **Musculoskeletal** | Non-significant | 0.005 | - |
| **Cancer** | Non-significant | 0.062 | - |
| **Respiratory** | Non-significant | 0.005 | - |

**Figure S1. CONSORT Flow Diagram of Enrollment Pathways.** The diagram illustrates the inclusion process for participants in the Wearable Enhancing All of us Research (WEAR) and Bring Your Own Device (BYOD) enrollment pathways. Wearable data included step counts and heart rate.

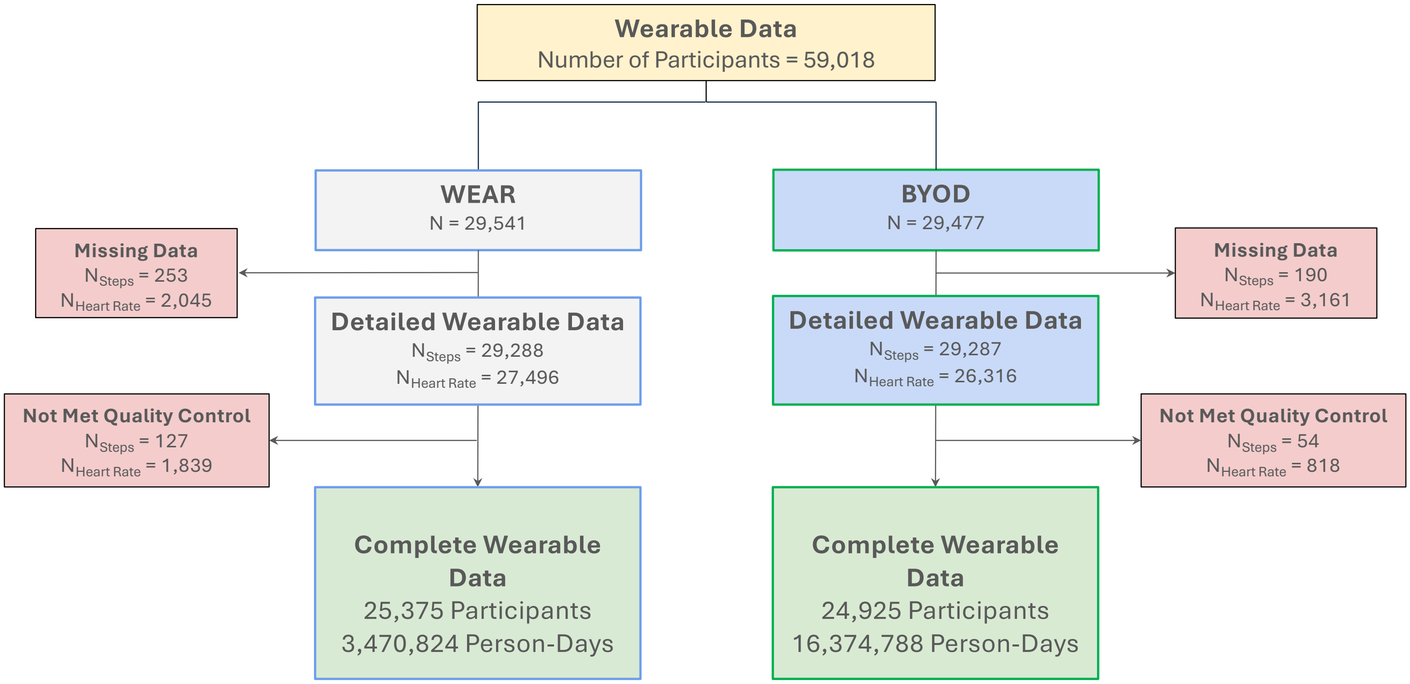

**Figure S2. Adherence to Wearable Monitoring Across Enrollment Pathways.** The graph represents the distribution of average daily wear time in the Wearable Enhancing All of Us Research (WEAR) and Bring Your Own Device (BYOD) pathways.

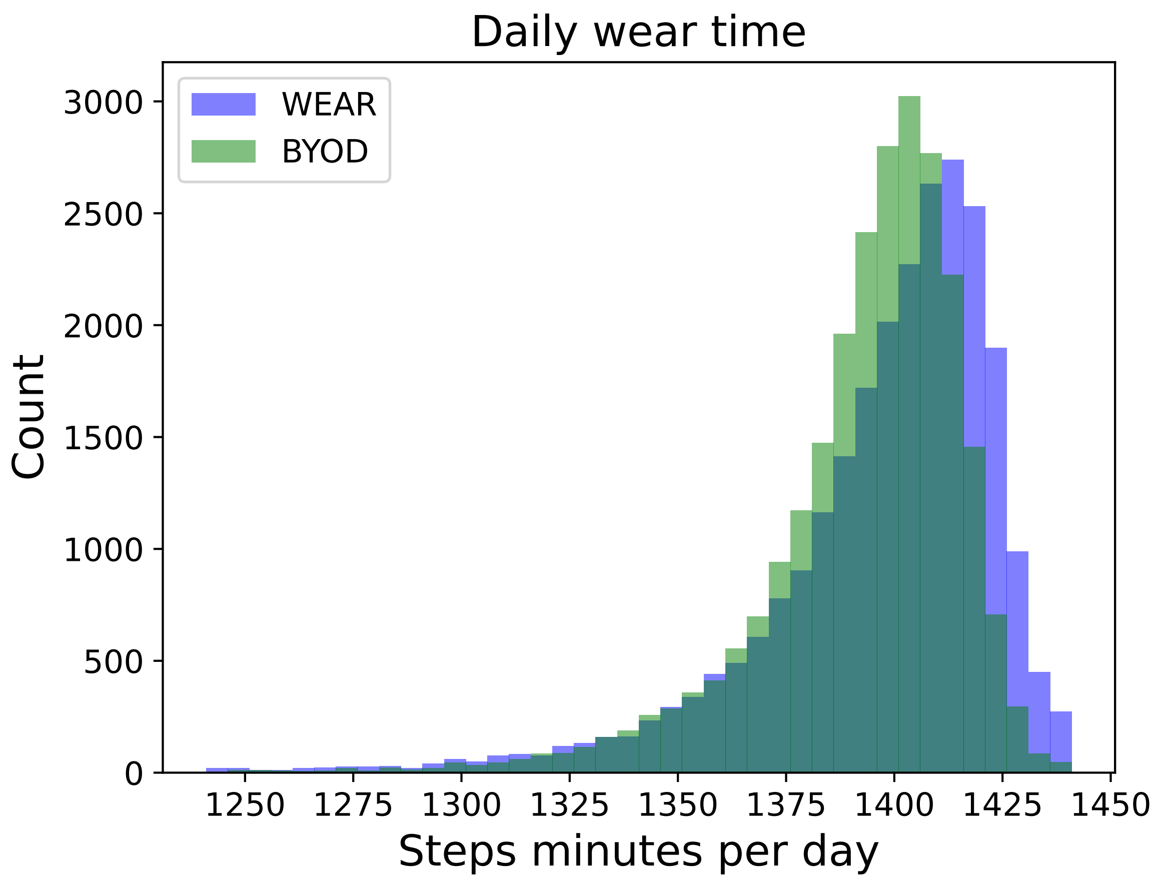

**Figure S3. Geographic Distribution of All of Us Research Program Participants.** State-level counts of participants are shown for the full All of Us Research Program cohort (A) and for the subset of participants with both Fitbit and electronic health records (EHRs) data available (B). Color scales represent the number of participants from each state, with yellow indicating higher and blue indicating lower counts. Abbreviations: EHR, electronic health records.

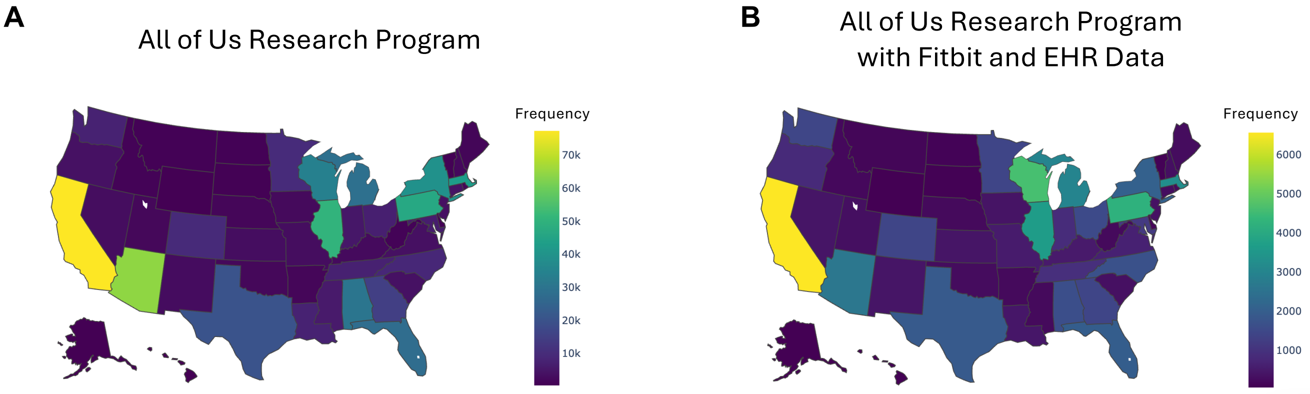

**Figure S4. Impact of COVID-19 Pandemic on Step Count.** Average daily step counts from 2018 to 2021 are shown across three periods: Pre-COVID (Jan 1, 2018–Jan 30, 2020), Lockdown (early 2020, shaded), and Post-lockdown (June 30, 2020–Dec 31, 2021). Each point represents a distinct month within this time interval.

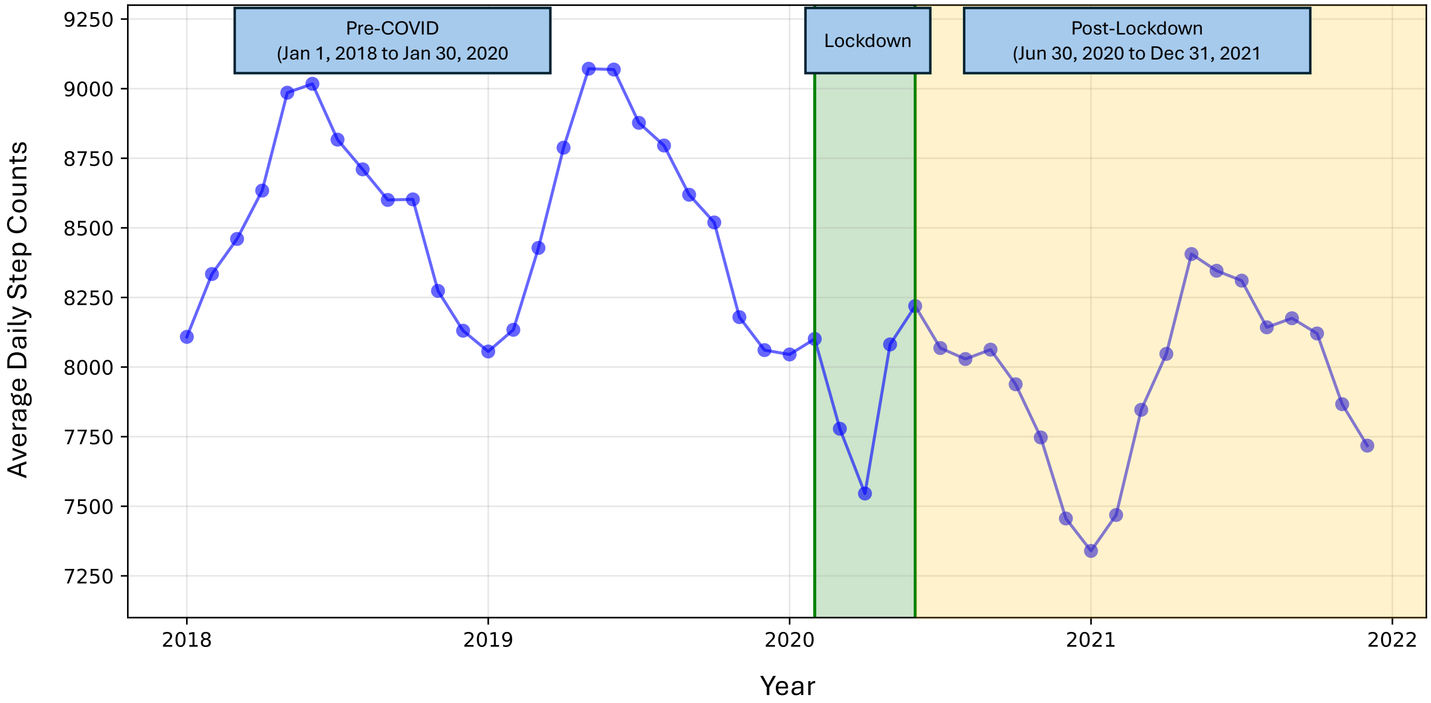

**Figure S5. Impact of Quality Control on Step Count Distribution.** Minute-level step count distributions shown before and after quality control procedures. The raw data (A) exhibit erratic fluctuations and wide confidence intervals due to artifacts and outliers. After applying quality control (B), the distribution shows a clear diurnal pattern with smooth estimates. Each line represents the mean step counts with 95% confidence intervals.

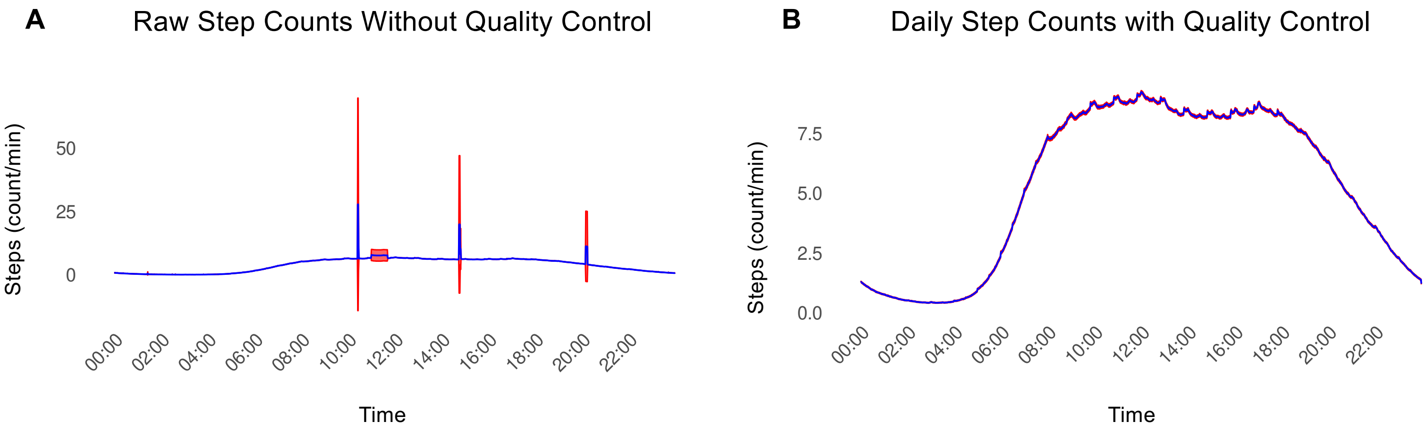

**Figure S6. Monthly and Multi-year Variations of Step Count and Sedentary Time.** Daily step counts (top) and sedentary times (bottom) are shown across monthly (left) and multi-year time scales (right). Plots represent the mean value at each time point with 95% confidence intervals. Multi-year analyses were restricted to individuals with at least 5 years of contiguous wearable data (n=3,743).

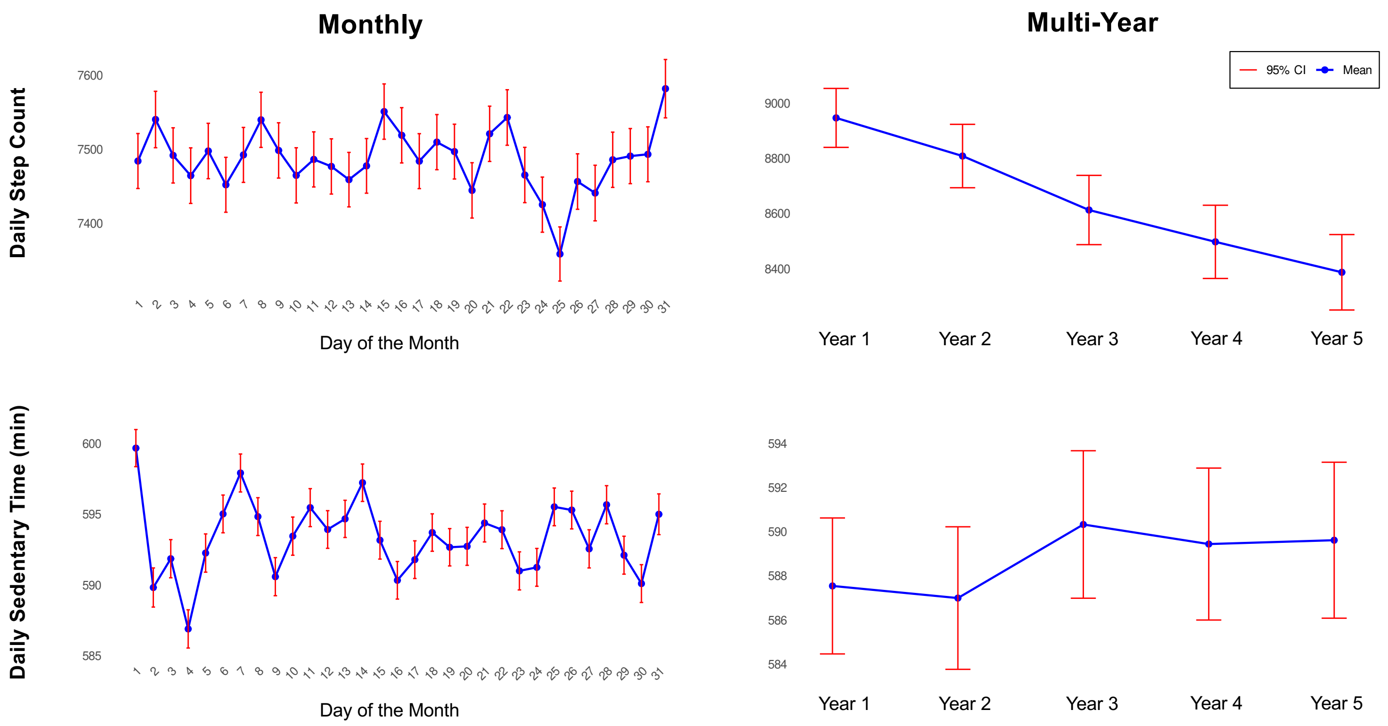

**Figure S7. Seasonal Variations of Step Count and Sedentary Time Across US States.** Graphs illustrate seasonal variations in daily average step count and sedentary time across all U.S. states and the District of Columbia. Color scales represent normalized distribution of each metric across states, with yellow indicating higher and blue indicating lower values.

A. Step Count

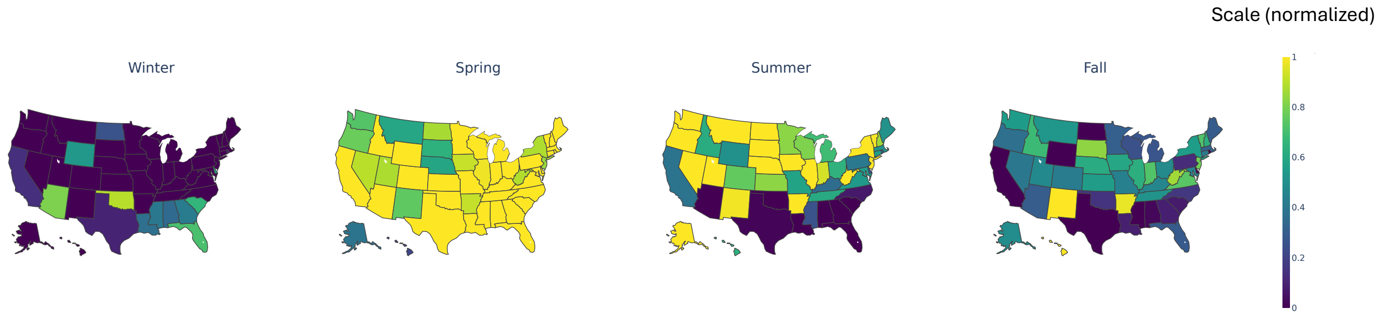


B. Sedentary Time

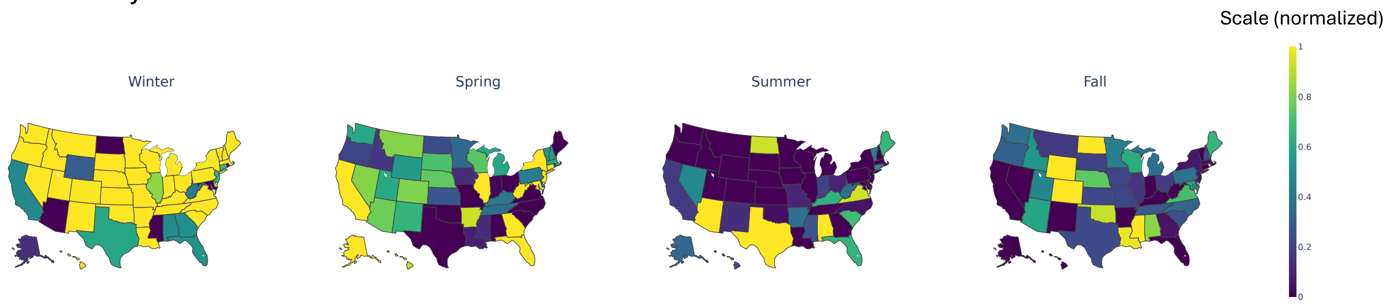

**Figure S8. Weekly Step Counts Across Sociodemographic Subgroups.** Weekly variations in step count are shown across sociodemographic subgroups of age, sex, self-reported race, education, income, and employment status. Bars represent average daily step counts with 95% confidence intervals.

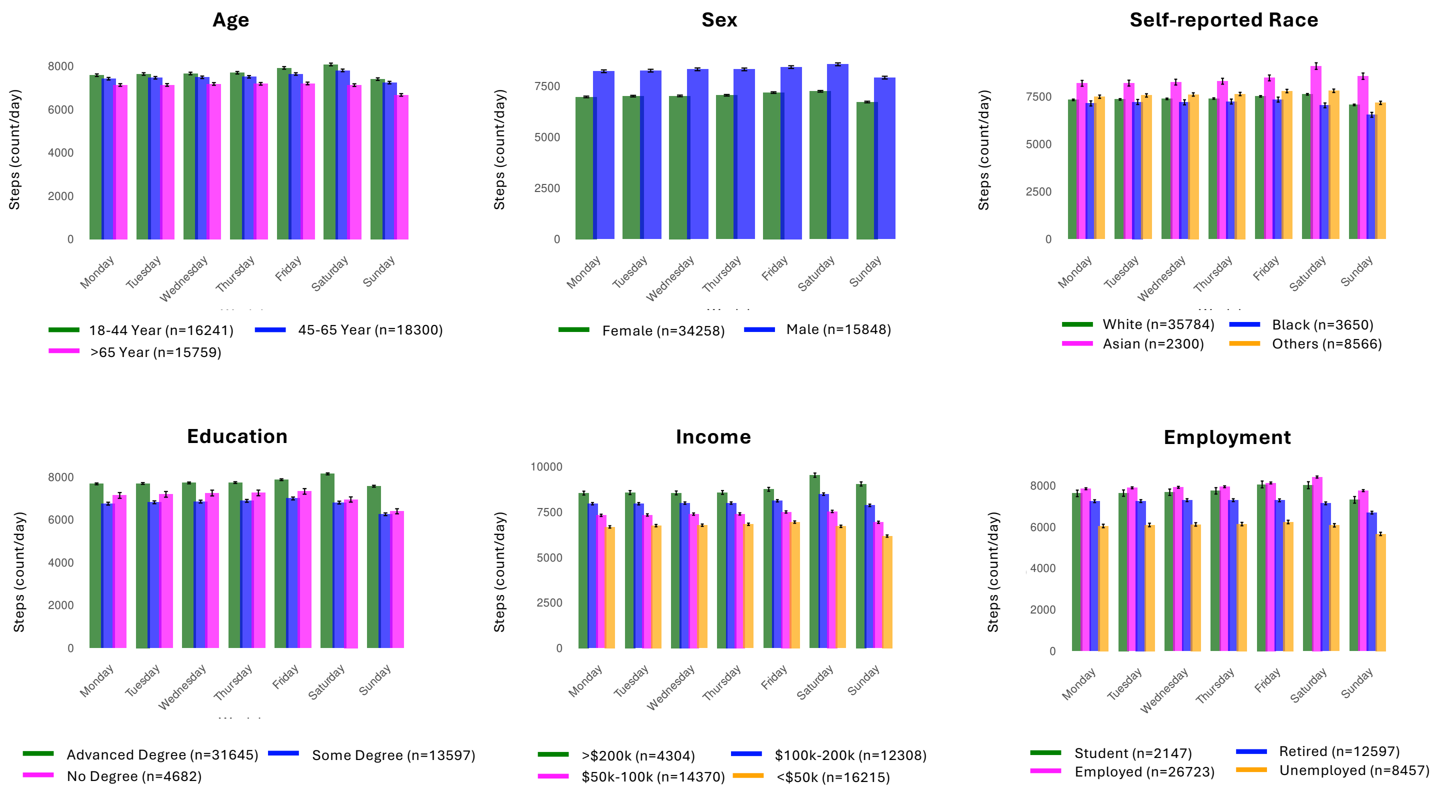

**Figure S9. Daily Step Counts in the BYOD Enrollment Pathway.** Minute-level time series of step counts across sociodemographic subgroups of age, sex, self-reported race, education, income, and employment status in the Bring Your Own Device (BYOD) pathway. Lines represent average step counts with 95% confidence interval.

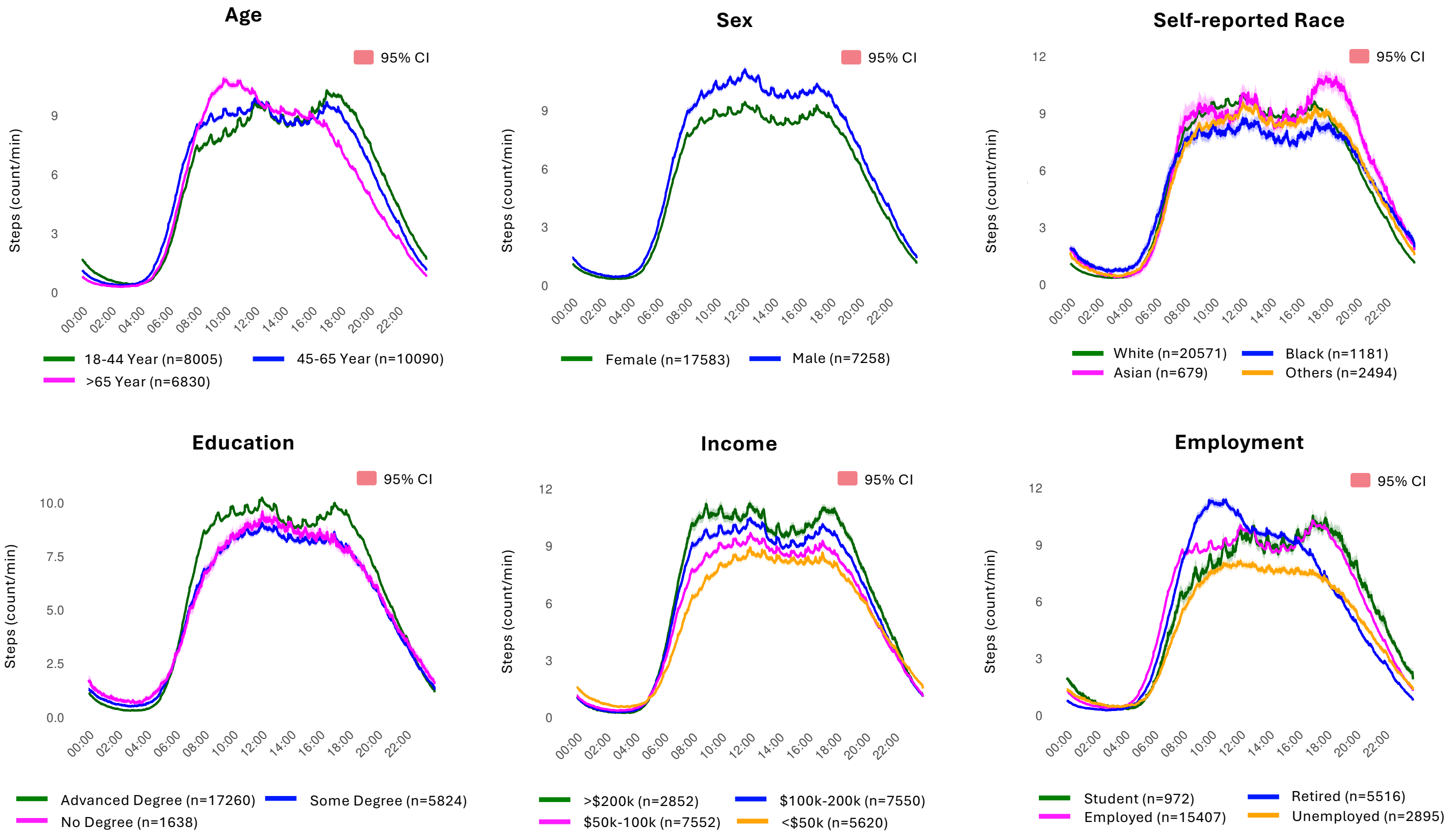

**Figure S10. Daily Step Counts in the WEAR Enrollment Pathway.** Minute-level time series of step counts across sociodemographic subgroups of age, sex, self-reported race, education, income, and employment status in the Wearable Enhancing All of us Research (WEAR) pathway. Wearables were provided at no cost to participants in the WEAR pathway. Lines represent average step counts with 95% confidence interval.

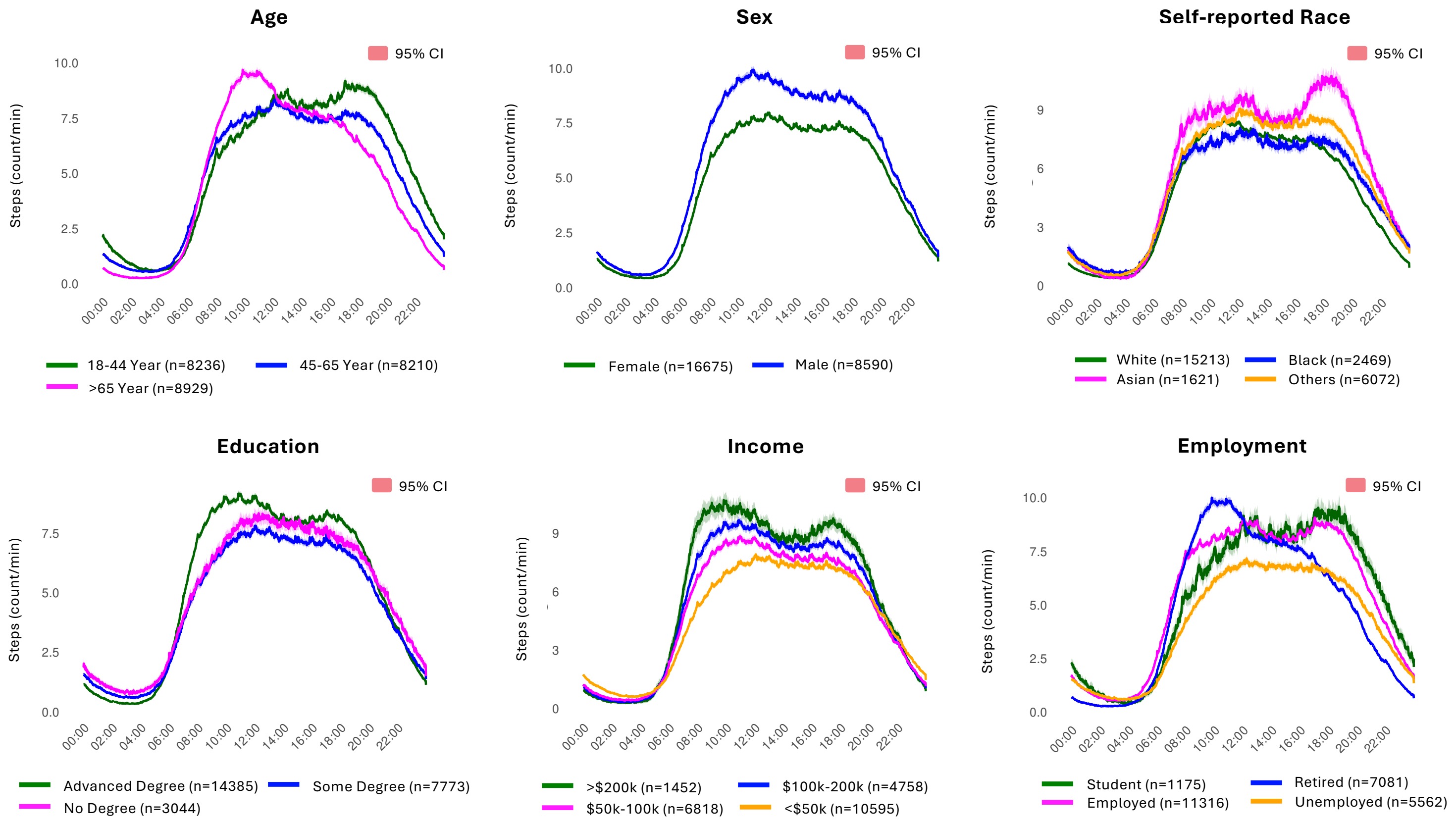

**Figure S11. Weekly Step Counts in the BYOD Enrollment Pathway.** Weekly patterns of step count across sociodemographic subgroups of age, sex, self-reported race, education, income, and employment status in the Bring Your Own Device (BYOD) pathway. Bars represent average daily step counts with 95% confidence intervals.

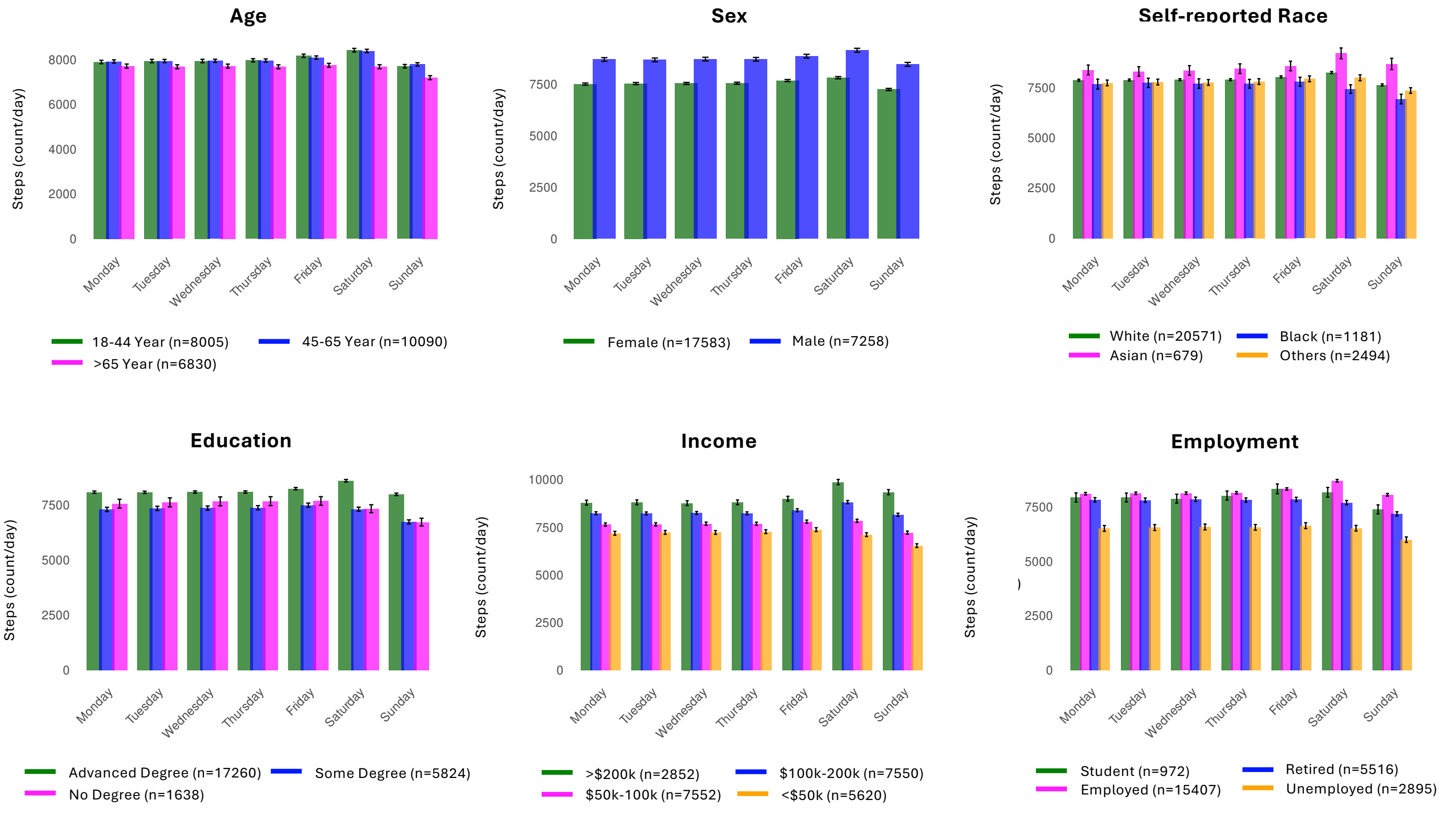

**Figure S12. Weekly Step Counts in the WEAR Enrollment Pathway.** Weekly patterns of step count across sociodemographic subgroups of age, sex, self-reported race, education, income, and employment status in the Wearable Enhancing All of us Research (WEAR) pathway. Wearables were provided at no cost to participants in the WEAR pathway. Bars represent average daily step counts with 95% confidence intervals.

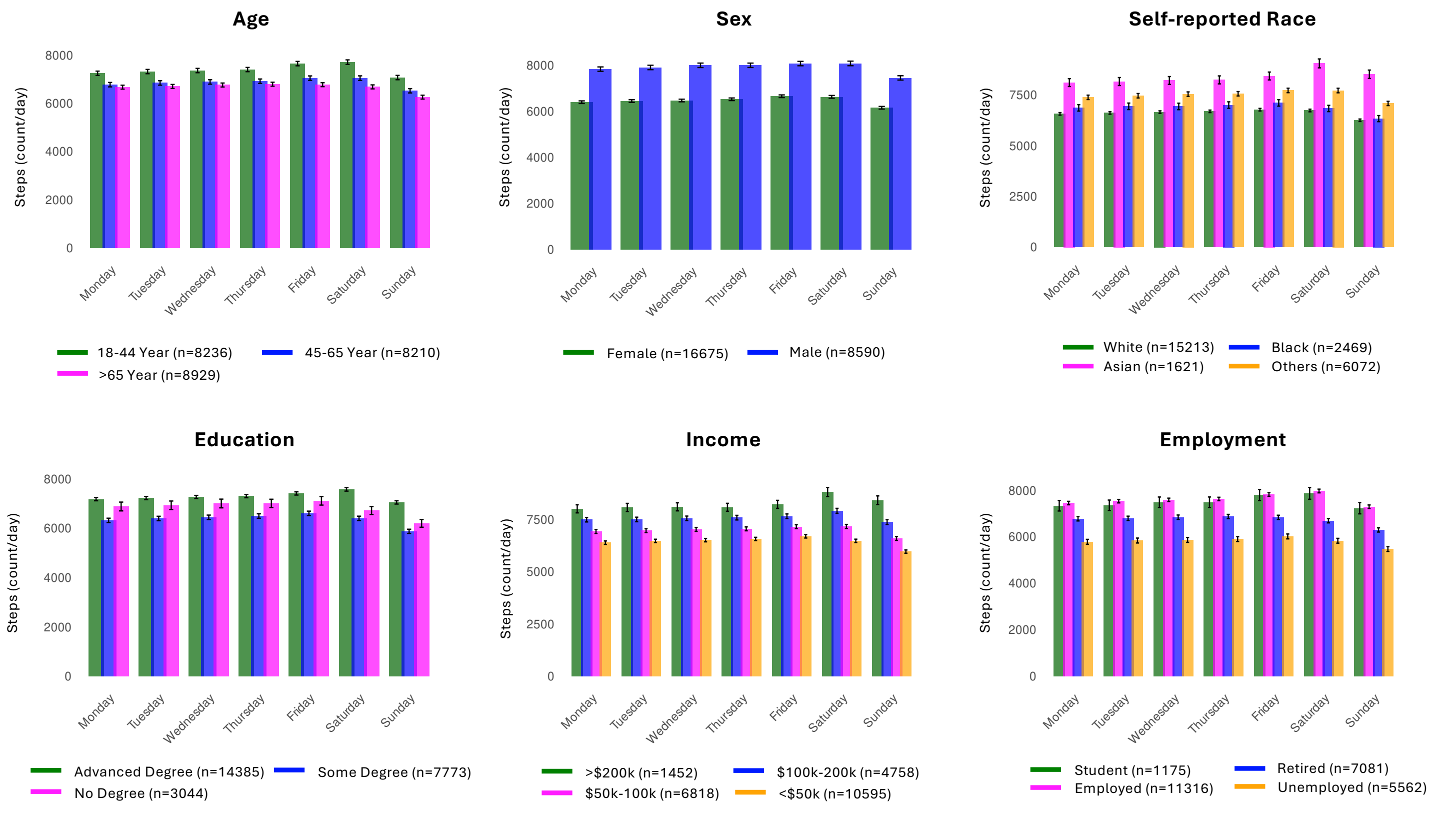

**Figure S13. Dose-Response Analysis of Daily Step Counts and Sedentary Time Deciles.** The figures represent cumulative incidence rate of eight major disease categories across deciles of daily step count (A) and sedentary time (B). Corresponding values of step count and sedentary time for each decile are shown.

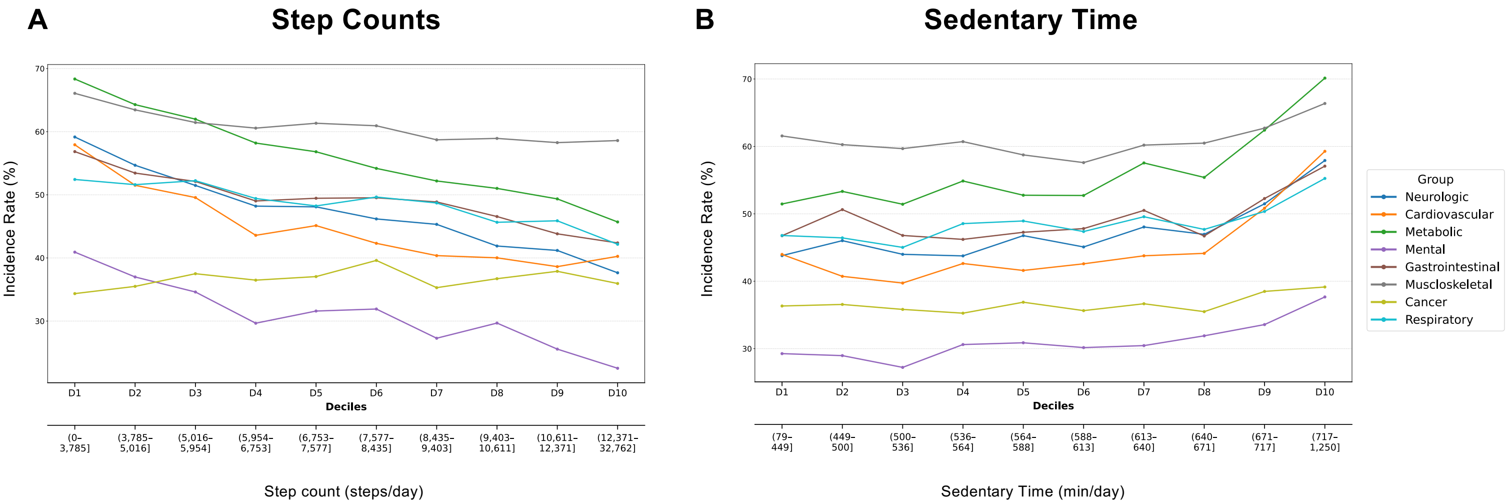
